# R1441G but not G201S mutation enhances LRRK2 mediated Rab10 phosphorylation in human peripheral blood neutrophils

**DOI:** 10.1101/2021.01.28.21249614

**Authors:** Ying Fan, Raja S. Nirujogi, Alicia Garrido, Javier Ruiz Martínez, Alberto Bergareche-Yarza, Elisabet Mondragón Rezola, Ana Vinagre Aragón, Ioana Croitoru, Ana Gorostidi Pagola, Laura Paternain Markinez, Roy Alcalay, Richard A. Hickman, Jonas Duering, Neringa Pratuseviciute, Shalini Padmanabhana, Francesc Valldeoriola, Maria José Martí, Eduardo Tolosa, Dario R Alessi, Esther Sammler

**Affiliations:** Medical Research Council Protein Phosphorylation and Ubiquitylation Unit, University of Dundee, Dundee, DD1 5EH, UK; Parkinson’s disease and Movement Disorders Unit, Neurology Service, Hospital Clínic de Barcelona, Barcelona, Spain; Centro de Investigación Biomédica en Red Sobre Enfermedades Neurodegenerativas (CIBERNED), Hospital Clínic, IDIBAPS, Universitat de Barcelona, Barcelona, Spain; Group of Neurodegenerative Diseases, Biodonostia Health Research Institute, San Sebastian, Spain; Department of Neurology, Columbia University Medical Center, New York, NY, USA; Department of Pathology and Cell Biology, Columbia University Medical Center New York, NY, USA; The Michael J Fox Foundation for Parkinson’s Research, New York, NY, USA; Molecular and Clinical Medicine, Ninewells Hopsital and Medical School, University of Dundee, DD1 9SY, UK

## Abstract

Gain-of kinase function variants in LRRK2 (leucine-rich repeat kinase 2) cause Parkinson’s disease (PD), albeit with incomplete and age-dependent penetrance, offering the prospect of disease-modifying treatment strategies via LRRK2 kinase inhibition. LRRK2 phosphorylates a subgroup of RabGTPases including Rab10 and pathogenic mutations enhance LRRK2-mediated phosphorylation of Rab10 at Thr73.

In this study we analyse LRRK2 dependent Rab10^Thr73^ phosphorylation in human peripheral blood neutrophils isolated from 101 individuals using quantitative immunoblotting and mass spectrometry. Our cohort includes 42 LRRK2 mutation carriers (21 with the G2019S mutation that resides in the kinase domain and 21 with the R1441G mutation that lies within the ROC-COR domain), 27 patients with idiopathic PD, and 32 controls.

We show that LRRK2 dependent Rab10 ^Thr73^ phosphorylation is significantly elevated in all R1441G LRRKR2 mutation carriers irrespective of disease status. PD manifesting and non-manifesting G2019S mutation carriers as well as idiopathic PD samples did not display elevated Rab10 ^Thr73^ phosphorylation. Furthermore, we analysed brain samples of 10 G2019S and 1 R1441H mutation carriers as well as 10 individuals with idiopathic PD and 10 controls. We find high variability for pRab10^Thr73^ phosphorylation amongst donors irrespective of genetic and disease state.

We conclude that in vivo LRRK2 dependent pRab10^Thr73^ analysis in human peripheral blood neutrophils is a specific and robust biomarker for LRRK2 kinase activation for individuals with mutations such as R1441G that enhance pRab10^Thr73^ phosphorylation over 2-fold. We provide the first evidence that the LRRK2 R1441G mutation enhances LRRK2 kinase activity in a primary human cell.

## Introduction

Parkinson’s disease (PD) is a common neurodegenerative condition that affects 1% of people over the age of 60 and over 6 million people worldwide (*1*). Patients with PD present a wide spectrum of progressive motor and non-motor symptoms reflecting the relentless loss of neurons and neuronal function that often predate clinical symptom onset by decades (*2*). The greatest unmet need in PD are disease modifying treatments that slow or stop disease progression which is further highlighted by the projected doubling of PD cases over the next 20 years (*1*). While the underlying cause for PD is largely unknown, the discovery of rare genetic forms of the condition has provided crucial insight into pathomechanistic processes altered in PD that have been leveraged for devising novel targeted treatment strategies(*3*).

The Leucine rich repeat kinase 2 (LRRK2) is such a highly persued therapeutic target. It is a large multidomain protein made up of 51 exons and 2537 amino acids with a predicted molecular weight of 286 kDaltons. Its catalytic core is made up of a kinase and ROC-COR GTPase domain as well as other protein-protein interaction motifs: the N-terminal armadillo, ankyrin and leucine-rich repeats motifs and a C-terminal WD40 motif (*4*). LRRK2 is highly expressed in immune cells including peripheral blood neutrophils and monocytes as well as lung, kidney and intestine but expression is lower in brain (*5, 6*). Pathogenic variants in LRRK2 are a direct cause for PD albeit with age-dependent and incomplete penetrance and cluster within the 2 catalytic domains (*7*). The substitution of serine for glycine at position 2019 within its kinase domain is the most common PD associated variant and accounts for 1% of sporadic and 4% of familial PD cases in most Caucasian populations worldwide, but up to 29% and 37% of familial cases in Ashkenazy Jews and North African Berbers, respectively, while being largely absent in Asian populations (*3, 8, 9*). The much rarer I2020T mutation is also located in the LRRK2 kinase domain. The second most common mutation hotspot is in the ROC-COR GTPase domain including R1441C/G/H/S as well as N1437H and Y1699C (*4*). Of these, the R1441G mutation is particularly common in the Basque region where it is responsible for 46% of all familial PD in this region of Spain (*10*). In addition to these clearly pathogenic mutations, genome wide association studies have implicated variants at the LRRK2 locus as risk factors for idiopathic PD with a modest increase in lifetime susceptibility for PD (*11, 12*).

All pathogenic LRRK2 mutations have in common that they augment kinase activity suggesting that inhibition of the LRRK2 kinase with small molecules is a promising strategy for disease modification. The LRRK2 kinase domain mutations – G2019S and I2020T – increase LRRK2 kinase activity only modestly, under 2-fold *in vitro* and *in vivo* cell and animal studies (reviewed in(*13*)) by domain disruption. The LRRK2 ROC and COR domain mutations – N1437H, R1441 and Y1699C – suppress GTPases activity and promote GTP binding which in turn mediates a 3-4-fold increase in LRRK2 kinase activity(*14, 15*). Assessing LRRK2 kinase activation status in human bio-samples on the other hand has been a challenge. For example, while LRRK2 autophosphorylation at Serine 1292 correlates with LRRK2 kinase activity, its low stoichiometry makes its detection difficult and unreliable. Serine 935 is one of the constitutively phosphorylated LRRK2 sites that reside in a non-catalytic region and plays a role in 14-3-3 binding (*16*). Serine 935 phosphorylation is one of the principal pharmacodynamic markers for *in vivo* LRRK2 kinase inhibition(*17*), but not for LRRK2 kinase activity. More recently, the discovery of a subgroup of RabGTPases, including Rab10, as endogenous LRRK2 kinase substrates that are phosphorylated at a conserved Threonine or Serine in conserved switch II domains (*14*), the availability of relevant tools (*5*) and technologies(*18*) as well as the exploration of novel biomatrices(*19*) has opened up new opportunities to assess how LRRK2 mutations impinge on is kinase activity.

We have previously described a robust and facile assay for interrogating LRRK2 kinase pathway activity in peripheral blood neutrophils by quantifying LRRK2 mediated phosphorylation of Rab10 at Thrronine 73 (*20, 21*). Phosphorylation of Rab10 at Thr73 is quantified by either immunoblot analysis deploying a highly sensitive monoclonal phosphospecific antibodies raised against the pRab10 Thr73 epitope(*20*) or by targeted mass-spectrometry approaches (*18*). As a biomatrix, peripheral blood neutrophils lend themselves for the study of the LRRK2 pathway as they constitute a homogenous and abundant pool of cells that contain relatively high protein copy numbers of both LRRK2 and Rab10 (*20, 21*). We have recently utilized this assay to demonstrate that LRRK2 dependent Rab10 phosphorylation is significantly increased in PD patients with a heterozygous VPS35 D620N mutation, that activates LRRK2 kinase activity by a yet unknown mechanism. To date elevated LRRK2 kinase activation has not been demonstrated in bio-samples from LRRK2 mutation carriers.

In this study, we have analysed LRRK2 dependent Rab10^Thr73^ phosphorylation as a marker for in vivo LRRK2 activation status in human peripheral blood neutrophils isolated from 101 individuals including 42 LRRK2 mutation carriers (21 with the G2019S mutation that resides in the kinase domain and 21 with the R1441G mutation that lies within the ROC-COR domain) with and without PD and compared them with 32 healthy controls and 27 individuals with idiopathic PD. We show that LRRK2 dependent Rab10 ^Thr73^ phosphorylation is significantly elevated in all R1441G LRRKR2 mutation carriers irrespective of disease status while PD manifesting and non-manifesting G2019S mutation carriers as well as idiopathic PD samples lacked any such enhancement over controls. We deployed two independent methodologies – quantitative multiplex immunoblotting for pRab10 normalized against total Rab10 as well as targeted pRab10^Thr73^ occupancy mass-spectrometry and found realatvely good correlation between the 2 assays. Furthermore, we have analysed brain samples of 10 G2019S and 1 R1441H mutation carriers as well as 10 individuals with idiopathic PD and 10 controls for LRRK2 dependent pRab10^Thr73^ phosphorylation and find high variability amongst donors irrespective of genetic and disease state.

## Materials and Methods

A comprehensive list of reagents, antibodies, cDNA constructs, mice, cell lines, buffers, equipment, software packages utilized in this study are provided in Supplementary table S1:

### Reagents

Cis-2,6-dimethyl-4-(6-(5-(1-methylcyclopropoxy)-1H-indazol-3-yl)pyrimidin-4-yl)morpholine (MLi-2) (*22, 23*) was synthesised at the University of Dundee. Diisopropylfluorophosphate(DIFP) was purchased from Sigma (Cat# D0879), Microcystin-LR was from Enzo Life Sciences and sequencing grade modified trypsin from Promega (Cat# V511A). Complete protease and phosphatase inhibitor tablets were from Roche. All heavy and light stable isotope synthetic peptides described in supplementary table 1 were synthesized by JPT peptide technologies (https://www.jpt.com/) in 1 nano mole aliquots. All the synthetic peptides were quantified by amino acid analysis and LC-MS analysis by JPT and confirmed to be of purity of >95%. The peptides were delivered in a lyophilized form and were resuspended in solvent containing 0.1% (v/v) formic acid in 3% (v/v) acetonitrile to give a final concentration of 10 pmol/μl. Aliquots of this were further diluted in a series of 10-fold dilution to a lowest concentration of 10 fmol/μl and stocks of each dilution aliquoted and stored at −80 °C.

**Table 1:**
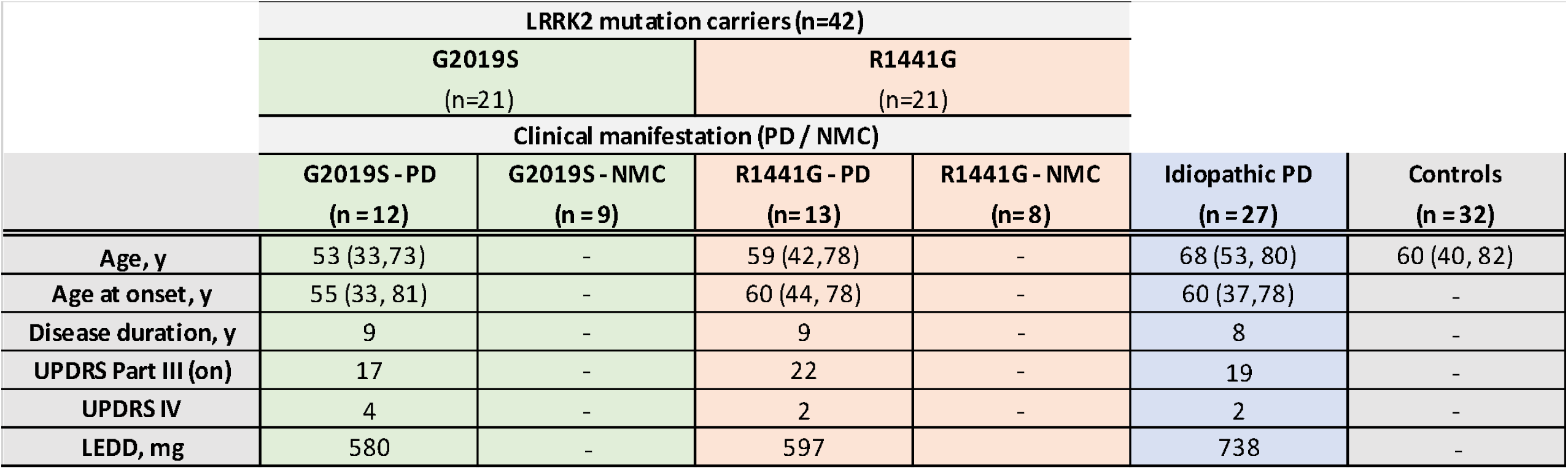
Summary of demographic and clinical characteristics of participants (n=101). Of the 101 participants, 42 carried either the LRRK2 kinase domain mutation G2019S or the ROC-COR domain mutation R1441G and were either diagnosed with PD or non-manifesting mutation carriers (NMC). Mean age in years (y) and age range in parenthesis, mean disease duration in years with range in parenthesis, *UPDRS* is Unified Parkinson’s Disease Rating Scale, part III (motor symptoms) in the on state and part IV (motor complications) as well as L-dopa equivalent daily dosage (LEDD) in mg.

|  | LRRK2 mutation carriers (n=42) |  |  |  | Idiopathic PD<br>(n = 27) | Controls<br>(n = 32) |
| --- | --- | --- | --- | --- | --- | --- |
|  | G2019S<br>(n=21) |  | R1441G<br>(n=21) |  |  |  |
|  | Clinical manifestation (PD / NMC) |  |  |  |  |  |
|  | G2019S - PD<br>(n = 12) | G2019S - NMC<br>(n = 9) | R1441G - PD<br>(n = 13) | R1441G - NMC<br>(n = 8) |  |  |
| Age, y | 53 (33,73) | - | 59 (42,78) | - | 68 (53, 80) | 60 (40, 82) |
| Age at onset, y | 55 (33, 81) | - | 60 (44, 78) | - | 60 (37,78) | - |
| Disease duration, y | 9 | - | 9 | - | 8 | - |
| UPDRS Part III (on) | 17 | - | 22 | - | 19 | - |
| UPDRS IV | 4 | - | 2 | - | 2 | - |
| LEDD, mg | 580 | - | 597 | - | 738 | - |

### Antibodies

The recombinant MJFF-pRab10 (Thr73) rabbit monoclonal antibodies were recently described in terms of their high selectivity and specificity (*5*) and are available from Abcam (Cat# ab230261). The MJFF-total Rab10 mouse antibody was from nanoTools (Cat# 0680–100/Rab10-605B11). Rabbit monoclonal antibodies for total LRRK2 (N-terminus, residues 100-500, UDD3) and phospho-Ser935 LRRK2 (UDD2) were expressed and purified at University of Dundee as described previously (*24, 25*) and are available from MRC PPU Reagents and Services (https://mrcppureagents.dundee.ac.uk/). The C-terminal total LRRK2 mouse monoclonal antibody was from Neuromab (Cat# 75-253). The phospho-Rab12 (S106) rabbit monoclonal antibody was described earlier(*26*). Mouse anti-GAPDH total was from Santa Cruz Biotechnology (Cat# sc-32233). The sheep polyclonal PPM1H antibodies aginst the full length PPM1H protein has been described before(*27*) and is available from MRC PPU Reagents and Services (sheep number DA018 https://mrcppureagents.dundee.ac.uk/). Recombinant antibodies were used at 1 mg/ml final concentration for immunoblotting except for anti-Rab10 (phospho T73) antibody which was used at 0.5 μg/ml final concentration. For immunoblotting applications all commercial monoclonal antibodies were diluted in 5% (w/v) bovine serum albumin in TBS-T (20 mM Tris base, 150 mM Sodium Chloride (NaCl), 0.1% (v/v) Tween20), and sheep polyclonal antibodies were diluted in 5% (w/v) skimmed milk in TBS-T. Goat anti-mouse IRDye 800CW (Cat# 926-32210) and IRDye 680LT (Cat# 926-68020), goat anti-rabbit IRDye 800CW (Cat# 926-32211) secondary antibodies were from LI-COR and used at 1:10000 dilution in TBS-T.

### Study participants and blood sample collection

Fresh blood was collected from a total of 101 participants; 66 individuals were recruited via the movement disorder clinics at Hospital Clinic Universitari de Barcelona in the fall of 2017 and 35 individuals via the Hospital Universitario Donostia in San Sebastian in the Basque region in Spain during 2019. Of the 101 participants, 42 carried a pathogenic mutation in LRRK2 −21 carried the G2019S mutation that resides in the kinase domain and 21 with the R1441G mutation that lies within the ROC-COR domain-, 27 patients with idiopathic PD, and 32 controls. Demographics such as gender, age, disease duration and age at PD onset were collected. Diagnosis of Parkinson’s was defined according to the UK Brain Bank criteria with the exception that a positive family history for PD was not considered an exclusion criteria (*28*). Severity of motor symptoms and the presence of motor complications was assessed using part III and IV of the Movement disorder society – Unified Parkinson’s disease rating scale (MDS-UPDRS-III, -IV) (*29*). Levodopa-equivalent daily dose (LEDD) was recorded as well as any additional non-oral therapies such as Deep Brain Stimulation (DBS). Detailed demographic and clinical information of all participants can be found in Supplementary Table S2.

### Ethical approval and consent to participate

The study was approved by the respective local ethics committees. All participants gave written informed consent.

### Neutrophil isolation, treatments and lysis

Peripheral blood neutrophils were isolated directly from fresh blood using EasySep Direct Human Neutrophil Isolation Kit (Stemcell Technologies, Cat# 19666) based on an immunonegative magnetic selection process as described before (*19, 20*). Pure and alive neutrophils were then pelleted by centrifugation at 335 g for 5 minutes, resuspended in 20ml RPMI 1640 media and divide equally into two tubes, one for LRRK2 kinase inhibitor treatment with MLi-2 at a final concentration of 200nM for 30 min and the other for vehicle control DMSO treatment. Neutrophils were then pelleted via centrifugation at 335 g for 5 minutes, the supernatant was carefully and fully removed before cell lysis in 150 μl of ice-cold lysis buffer containing 50 mM Tris/HCl, pH 7.5, 1% (v/v) Triton X-100, 1 mM ethylene glycol-bis(β-aminoethyl ether)-N,N,N⍰,N⍰-tetraacetic acid (EGTA), 1 mM sodium orthovanadate, 50 mM natrium fluoride (NaF), 0.1% (v/v) 2-mercaptoethanol, 10 mM 2-glycerophosphate, 5 mM sodium pyrophosphate, 1 μg/ml mycrocystin-LR (Enzo Life Sciences), 270 mM sucrose, 0.5 mM diisopropylfluorophosphate (DIFP) (Sigma, Cat# D0879) in addition to Complete EDTA-free protease inhibitor cocktail (Roche, Cat# 11836170001). Cell lysates were kept on ice for 10 min and then clarify by centrifugation at 20800 g for 15 min at 4 °C. Supernatants were used for Bradford assay (Thermo Scientific) and stored at −80 °C after snap freezing.

### Brain sample preparation

Brain samples were obtained from institutionally approved autopsy collections held by the Columbia University Medical Center in New York, USA and the IDIBAPS Biobank at the Hospital Clinic in Barcelona, Spain.

Frozen human matched frontal (Brodmann area 9) and occipital (Brodmann area 17) cortex samples from 10 individuals including 3 controls, 4 G2019S mutation carriers with PD and 3 iPD were obtained from the brain bank at the Columbia University Medical Center in New York. Additionally, we received 20 frontal cortex samples including 7 controls, 7 iPD, 5 G2019S and 1 R1441H mutation carriers with PD from the IDIBAPS Biobank at the Hospital Clinic in Barcelona, Spain. Brain samples were weighed and added to a 10-fold volume excess of ice-cold lysis buffer containing 50 mM Tris– HCl pH 7.5, 1% (v/v) Triton X-100, 1 mM EGTA, 1 mM sodium orthovanadate, 50 mM sodium fluoride, 10 mM β-glycerophosphate, 5 mM sodium pyrophosphate, 1 µg/ml microcystin-LR (Enzo Life Sciences), 270 mM sucrose and complete EDTA-free protease inhibitor cocktail (Roche, Cat # 11836170001), and homogenised using POLYTRON homogenizer (KINEMATICA) on ice (5 s homogenisation, 10 s interval and 5 s homogenisation). Lysates were cleared by centrifugation at 201.800 *g* for 10 min at 4 °C. Supernatants were collected, quantified by the Bradford assay (Thermo Scientific) and subjected to immunoblot analysis.

### Quantitative immunoblot analysis

Cell lysates were mixed with 4× SDS–PAGE loading buffer [250 mM Tris–HCl, pH 6.8, 8% (w/v) SDS, 40% (v/v) glycerol, 0.02% (w/v) Bromophenol Blue and 4% (v/v) 2-mercaptoethanol] to a final total protein concentration of 1 µg/µl and heated at 70°C for 10 min. Ten microgram of samples were loaded onto NuPAGE 4–12% Bis–Tris Midi Gel (Thermo Fisher Scientific, Cat# WG1403BOX) and electrophoresed at 130 V for 2 h with the NuPAGE MOPS SDS running buffer (Thermo Fisher Scientific, Cat# NP0001-02). At the end of electrophoresis, proteins were electrophoretically transferred onto the nitrocellulose membrane (GE Healthcare, Amersham Protran Supported 0.45 µm NC) at 100 V for 90 min on ice in the transfer buffer (48 mM Tris–HCl and 39 mM glycine). Transferred membrane was blocked with 5% (w/v) skim milk powder dissolved in TBS-T [20 mM Tris–HCl, pH 7.5, 150 mM NaCl and 0.1% (v/v) Tween 20] at room temperature for 1 h. The membrane was then cropped into three pieces, namely the ‘top piece’ (from the top of the membrane to 75 kDa), the ‘middle piece’ (between 75 and 30 kDa) and the ‘bottom piece’ (from 30 kDa to the bottom of the membrane). The top piece was incubated with rabbit anti-LRRK2 pS935 UDD2 antibody multiplexed with mouse anti-LRRK2 C-terminus total antibody diluted in 5% (w/v) skim milk powder in TBS-T to a final concentration of 1 µg/ml for each of the antibody. The middle piece was incubated with mouse anti-GAPDH antibody diluted in 5% (w/v) skim milk powder in TBS-T to a final concentration of 50 ng/ml. The bottom pieces were incubated with rabbit MJFF-pRAB10^clone-1^ monoclonal antibody multiplexed with mouse MJFF-total Rab10^clone-1^ monoclonal antibody diluted in 2% (w/v) bovine serum albumin in TBS-T to a final concentration of 0.5 µg/ml for each of the antibody. All blots were incubated in primary antibody overnight at 4°C. Prior to secondary antibody incubation, membranes were washed three times with TBS-T for 10 min each. The top and bottom pieces were incubated with goat anti-mouse IRDye 680LT (#926-68020) secondary antibody multiplexed with goat anti-rabbit IRDye 800CW ((#926-32211) secondary antibody diluted in TBS-T (1⍰:⍰10⍰000 dilution) for 1 h at room temperature. The middle piece was incubated with goat anti-mouse IRDye 800CW (#926-32210) secondary antibody diluted in TBS-T (1⍰:⍰10⍰000 dilution) at room temperature for 1 h. Membranes were washed with TBS-T for three times with a 10 min incubation for each wash.

### Quantification and normalization of the LICOR immunoblot analysis

Quantification of the protein bands was performed on the scanned images using the Odyssey Scan band tool in a blinded manner with regards to genotype and clinical status of the participants by an independent analyst using the Image Studio software. Each sample set including DMSO and MLi-2 treated neutrophil lysates for each participant or each autospsy brain lysate was run in duplicates and 2 independent replicate immunoblotting experiments were performed and used for quantification. The intensities of the total LRRK2 and total Rab10 protein bands were normalized to that of the housekeeping protein GAPDH (total LRRK2/GAPDH and total Rab10/GAPDH) while the specific posttranslational phosphorylation modifications were quantified against the multiplexed total target protein irrespective of modification (Thr73-pRab10 /total Rab10 and Ser935-pLRRK2/total LRRK2). Inter-gel variability was controlled for by normalization against the same control sample on each gel per set of experiments. In total, 3 sets of Western blot experiments run with samples 1-66 (Barcelona set), samples 67-82 (San Sebastian set 1) and 83-101 (San Sebastian set 2) were performed at different time points. In order to combine Western blot results for all 3 sets of neutrophil lystaes, results for each set were normalized to the average of its controls and then combined.

### SDS-PAGE separation and In-gel digestion

The method utilized here was adopted according to the recently published paper by Karayel et al.(*18*). Briefly, neutrophil lysates were reduced with 5 mM dithiothreitol at 55 °C for 20 min. Samples were brought to room temperature and treated with 40 mM iodoacetamide for 20 min in the dark. 40 μg of total soluble protein extract of neutrophils were separated by SDS-PAGE and the gels were stained with colloidal Coomassie blue (Novex). Protein bands spanning 20 to 30 kDa region were excised, destained with 40% acetonitrile in 40 mM ammonium bicarbonate on a Thermomixer for 20 min at room temperature. Discard destaining solution and repeat the step once until gel pieces were completely destained. Gel pieces were dehydrated with 100% acetonitrile on a Thermomixer for 15 min, followed by complete removal and placing of the tubes on ice. The dried gel pieces were reswollen in 150 μl of 20 mM TEABC pH7.5 containing 500 ng sequencing grade modified trypsin and 0.1% sodium deoxycholate. Samples were incubated at 37 °C for 16 h. Peptides were extracted by adding to samples 70 μl of 0.5% acetic acid in 80% acetonitrile and incubate on a shaker for 15 min at ambient temperature. The liquid containing the peptide extraction was then transferred into a fresh tube. The extraction step was repeated twice with pooling of the peptide solutions with that of the previous step. Peptide solutions were snap frozen with dry ice and dried in a Speedvac evaporator.

### LC-MS/MS sample preparation

#### SDB-RP (Styrenedivinylbenzene-Reversed Phase) purification

SDB-RP tips were prepared in-house by punching two layers of 16 guage syringe needle and pushed into 250 ul of pipette tip. Vacumm dried peptides from the previous step were reconstituted with 100 µl of 1% trifluoroacetic acid in isopropanol (vol/vol) and directly loaded on SDB-RP tips. Samples were centrifuged at 1500 g at room temperature for 10 min to allow binding of peptides onto the discs. Flow-through was collected and reloaded onto stage tips for another round of binding. Subsequently, stage tips were washed twice with 100 ul of 1% trifluoroacetic acid in Isopropanol (vol/vol), and then by adding 100 ul of 0.2% Trifluroacetic acid in 3% Acetonitrile (vol/vol). Wash steps were undertaken by centrifugation at 2000 g for 6 minutes at room temperature and the flow-through was dicarded. Peptides were eluted sequentially with freshly prepared 1.25% ammonium hydroxide in 50% acetonitrile (vol/vol) and 1.25% ammonium hydroxide in 80% acetonitrile (vol/vol). Purified peptides were snap frozen with dry ice and dried in a Speedvac evaporator.

#### Spike in of heavy labeled paptides and sample loading on EvoTips

Sample loading on Evotips was performed as described previously (*27*) with a minor modififications as described below. Vacuum dried peptides were reconstituted in 80 ul of 0.1% Formic acid in 3% acetonitrile (vol/vol) buffer and allowed to dissolve at room temperature for 10 minutes. To calculate absolute phosphorylated Rab10^THr73^ occupancy, an equimolare mixture of 25 fmol of heavy stable isotope labeled (SIL) phosphorylated and non-phosphorylated Rab10 counterpart peptides and 25 fmol of PRTC retention time calibration mix were spiked in. The peptide sample mix was then subjected to sonication on a waterbath sonicator (Branson) for 10 minutes and centrifuged at 17,000g at room temperature for 10 minutes. Evotips (EvoSep, Cat #EV2001) were activated by adding 20 ul of buffer B (100% acetonitrile in 0.1% formic acid vol/vol) and centrifuged at 800g for a minute. Tips were immersed into 200ul of Isopropanol and then equilibrated by adding 20 ul of buffer A (0.1% formic acid in water vol/vol)) and centrifuged at 800g for a minute and repeated this step again. Peptidese were loaded on Evotips using gel loader tips and centrifuged at 700g for 5 minutes. The flowthrough was collected and reapplied and centfiguged. Next, the tips were washed by adding 0.1% buffer A and the washing was repeated again. A 100ul of buffer A was applied to the tips prior placing the Evotips on Autosampler tray of EvoSep LC system.

#### Parallel Reaction monitoring (PRM) measurements

All of the targeted LC-MS/MS (PRM) data was acquired on QE HF-X Mass spectrometer interfaced inline with an EvoSep liquid chromatrography system. EvoTips were loaded on EvoSep LC system autosampler tray and analyzed using 21 min (60 samples per day) script. The EvoSep LC system elutes peptides by applying a partial gradient using low pressure pump-A and B into the long storage capillary sample loop were peptides were partially stored and focused by pre-formed gradient using low pressure pump-C and pump-D, subsequently peptides were directed to an analytical column (Reprsoil-pur C18 AQ, 3um beads, 100um ID, 8cm long for pRab analysis) by high pressure pump which is housed into the Easy nano source. Peptides were then directly electrosprayed into the mass spectrometer by maintaining 2000 voltage. Mass spectrometer was operated in a targeted PRM mode by employing one Full MS scan followed by a PRM scan which is instructed to operate by following the imported inclusion list with a scheduled retention time (Peptide sequences and m/z values are provided in supplementary table 1). Dynamic on-the-fly retention time correction was enabled to correct the scheduled retention time. Full MS Scan was acquired within 300-800 m/z and acquired at 120,000 resolution m/z 200 using Orbitrap mass analyzer. Each of the targeted analyte was isolated using Quadrupole mass filter with a 0.7Da isolation window and fragmented using normalized 27% higher energy collisional dissociation (HCD) and measured at 30,000 resolution at 200 m/z and measured using Orbitrap mass analyzer. The loop cycle was maintained to 10 scan per duty cycle. The AGC targets for Full MS and PRM were at 3E6 and 1E5 ions respectively and a maximum of 50ms for MS1 and 300ms for PRM scan. For Limit of detection (LOD) experiments, the heavy pRab10 peptide was kept at constant amount 50fmol and the light pRab10 peptide varied from (0.01,0.1, 1, 10,50 and 200fmol) was directly spiked into 50ng Hela cell tryptic digest. PRM data was acquired as described above except the ion injection times for 0.01, 0.1 and 1 fmol data was maintained at 500ms.

#### Spectral library generation

The retention times of heavy stable peptides were determined by using an equimolar mixture of 50fmol and 25fmol of PRTC peptide mixture was acquired using Data dependent acquisition (DDA) mode on QE HF-X mass spectrometer in line with EvoSep LC system. The DDA data was acquired using one Full MS and 10 top N MS2 scans by disabling dynamic exclusion option. MS1 and MS2 scans were acquired at 120,000 and 30,000 resolution at 200 m/z and measured using Orbitrap mass analyser. Raw data was processed using Protoeme Disoverer 2.2 version using Sequest search algorithm against Uniprot Human proteome database that was appended with PRTC peptide sequences. Mass error tolerance was set for MS1 at 10ppm and 0.05Da for MS2. Carbamidomethylation of Cys as a fixed modification and Heavy lable (13C6 15N4) for Arg and 13C6 15N2) for Lys residues, phosphorylation of Ser/Thr were set as dynamic modification. Percolator node was used to filter that data at 1% FDR. The output .msf files were directly imported into Skyline software suite to serve as a spectral library which is then used for all of the subsequent Human neutrophils PRM data.

#### Data analysis and phosphorylation stoichiometry calculations

All of the PRM raw data was imported into Skyline software suite. In-house generated Spectral library was used to pick the precursor and top six fragement ions. Both doubly(z=2) and triple (z=3) charge states were used. Extrated ion cchromatrograms of both MS1 and MS2 fragemtn transitions were manually examined and adjusted for any interfering ions. The light/heavy internal standard ratio values were exported to calculate the pRab10 stoichiometry as described in (PMID: Karayle et.al). Brieefly, the pRab10 stoichiometry was calculated by taking the ratio of total amount of phosphorylation to the toal amount of both phosphorylated and non-phophorylated ratio that was represented in terms of percentage, [pRab10 L/H / pRab10 L/H+npRab10 L/H*100]. All of the Neutrophils samples phosphorylation stoichiometry calculations are provided in table 2.

**Table 2:** Quantification of Rab10^Thr73^ phosphorylation levels by immunoblotting and targeted mass-spectrometry. (A) Descriptive analysis of multiplexed immunoblotting of pRab10/total Rab10 ratio normalized to healthy control DMSO neutrophil samples of at least 2 independent experiments with duplicate loading (≥4 data points per sample) and targeted mass-spectrometry measuring absolute Rab10^Thr73^ occupancy (%) of 2 independent measurements including mean, standard deviation (SD), standard error of the mean (SEM), minimum and maximum for control, iPD and G2019S and R1441G mutation carriers. Segregation of LRRK2 mutation carriers according to clinical status did not reveal a statistical difference between mutation carriers with and without PD (p* of unpaired t-test (PD vs NMC). (B) One-way ANOVA followed by multiple comparisons where the mean of each column was compared against the means of all other groups with significance levels and adjusted p values for DMSO and MLi-2 treated samples. ns = non significant.

|  | Control DMSO | Control MLI-2 | iPD DMSO | iPD MLI-2 | G2019S DMSO | G2019S MLI-2 | R1441G DMSO | R1441G MLI-2 |
| --- | --- | --- | --- | --- | --- | --- | --- | --- |
| Multiplex immunoblotting<br>normalized pRab10/total Rab10 (DMSO) | (n=32) | (n=32) | (n=26) | (n=26) | (n=26) | (n=21) | (n=20) | (n=20) |
| Mean (ratio) | 1,00 | 1,00 | 0,94 | 0,96 | 1,06 | 1,08 | 4,61 | 0,99 |
| SD | 0,40 | 0,35 | 0,37 | 0,29 | 0,20 | 0,68 | 1,70 | 0,22 |
| SEM | 0,07 | 0,06 | 0,07 | 0,06 | 0,04 | 0,15 | 0,38 | 0,05 |
| Minimum | 0,11 | 0,46 | 0,21 | 0,42 | 0,53 | 0,41 | 1,80 | 0,70 |
| Maximum | 2,01 | 2,16 | 1,78 | 1,56 | 1,58 | 3,65 | 8,16 | 1,39 |
| Targeted mass-spectrometry<br>pRab10 (DMSO) Occupancy | (n=32) | (n=32) | (n=26) | (n=25) | (n=21) | (n=21) | (n=20) | (n=21) |
| Mean (%) | 1,52 | 0,30 | 1,61 | 0,25 | 1,90 | 0,28 | 6,60 | 0,27 |
| SD | 0,67 | 0,23 | 0,65 | 0,08 | 0,40 | 0,09 | 2,39 | 0,08 |
| SEM | 0,12 | 0,04 | 0,13 | 0,02 | 0,09 | 0,02 | 0,53 | 0,02 |
| Minimum | 0,20 | 0,10 | 0,20 | 0,15 | 1,01 | 0,18 | 2,85 | 0,20 |
| Maximum | 3,21 | 1,40 | 3,22 | 0,46 | 2,61 | 0,49 | 12,90 | 0,53 |

B
|  | Multiplex immunoblotting: normalized pRab10/total Rab10 |  |  |  | Targeted mass-spectrometry: pRab10 Occupancy (%) |  |  |  |
| --- | --- | --- | --- | --- | --- | --- | --- | --- |
| Tukey's multiple comparisons test | Summary | Adjusted P Value | Mean Difference | 95,00% CI of diff. | Summary | Adjusted P Value | Mean Difference | 95,00% CI of diff. |
| Control DMSO vs. iPD DMSO | ns | >0,9999 | 0,06 | -0,4656 to 0,5851 | ns | >0,9999 | -0,0872 | -0,7808 to 0,6064 |
| Control DMSO vs. G2019S DMSO | ns | >0,9999 | -0,06 | -0,6140 to 0,5036 | ns | 0,7636 | -0,3795 | -1,117 to 0,3583 |
| Control DMSO vs. R1441G DMSO | **** | <0,0001 | -3,61 | -4,177 to -3,043 | **** | <0,0001 | -5,073 | -5,822 to -4,325 |
| iPD DMSO vs. G2019S DMSO | ns | 1,00 | -0,11 | -0,6987 to 0,4689 | ns | 0,9414 | -0,2923 | -1,063 to 0,4785 |
| iPD DMSO vs. R1441G DMSO | **** | <0,0001 | -3,67 | -4,262 to -3,078 | **** | <0,0001 | -4,986 | -5,768 to -4,205 |
| G2019S DMSO vs. R1441G DMSO | **** | <0,0001 | -3,56 | -4,177 to -2,933 | **** | <0,0001 | -4,694 | -5,515 to -3,873 |
| Control DMSO vs. Control MLI-2 | ns | >0,9999 | 0,00 | -0,4975 to 0,4975 | **** | <0,0001 | 1,224 | 0,5669 to 1,880 |
| iPD DMSO vs. iPD MLI-2 | ns | >0,9999 | -0,02 | -0,5675 to 0,5362 | **** | <0,0001 | 1,361 | 0,6251 to 2,097 |
| G2019S DMSO vs. G2019S MLI-2 | ns | >0,9999 | -0,03 | -0,6421 to 0,5861 | **** | <0,0001 | 1,626 | 0,8158 to 2,437 |
| R1441G DMSO vs. R1441G MLI-2 | **** | <0,0001 | 3,62 | 2,994 to 4,252 | **** | <0,0001 | 6,331 | 5,490 to 7,173 |

#### Statistical analysis

Bioinformatic analyses in this study were performed with Skyline (https://skyline.ms/project/home/software/Skyline/begin.view), Microsoft Excel and Graphpad (GraphPad Software). Data visualization was done using GraphPad Prism (GraphPad Software). In general, grouped data analysis was performed using one-way ANOVA for multiple comparisons or unpaired t-test and displayed graphically using GraphPad Prism software (version 9).

## Results

### Clinical cohort and peripheral blood collection

A total of 101 participants were recruited, 66 individuals via the movement disorder clinics at Hospital Clinic Universitari de Barcelona in the fall of 2017 and 35 individuals via the Hospital Universitario Donostia in San Sebastian in the Basque region in Spain during 2019. Of the 101 participants, 42 carried a pathogenic mutation in LRRK2 −21 carried the G2019S mutation that resides in the kinase domain and 21 with the R1441G mutation that lies within the ROC-COR domain-, 27 patients with idiopathic PD, and 32 controls (Table 1). Amongst the LRRK2 mutation carriers were individuals with and without PD. In keeping with the age dependent penetrance of LRRK2, non-manifesting mutation carriers tended to younger than those with PD. All participants were clinically evaluated for PD motor symptoms and motor complications using the Unified Parkinson’s Disease Rating Scale (UPDRS) parts III and IV and age at onset of PD and PD duration as well as L-dopa equivalent daily dose (LEDD) calculated where applicable and age range (Supplementary table S2). 20 ml of fresh peripheral blood was collected for immediate neutrophil isolation followed by ex vivo neutrophil treatment with and without the specific LRRK2 kinase inhibitor MLi-2 prior to cell lysis and snap freezing for storage at −80 degrees Celsius.

All procedures were performed in compliance with the local ethics review boards and all participants provided informed consent.

### Experimental design and workflow

The experimental design and statistical rational for each experiment are described in each subsection and figure legend. Briefly, all peripheral blood neutrophil samples isolated from participants were subjected to duplicate quantitative multiplex immunoblotting as well as mass-spectrometry with technical replicates per analysis (≥2) (Figure 1). Analysis was performed blinded – in case of the quantitative immunoblotting by independent analysis and in case of the mass-spectrometry with genotype and clinical status only revealed after the mass-spectrometry measurements were completed. To ensure that any effect observed on the phosphorylation level of Rab10 at the Threonine 73 epitope was LRRK2 mediated, neutrophils were treated with and without the specific LRRK2 kinase inhibitor MLi-2.

**Figure 1:**
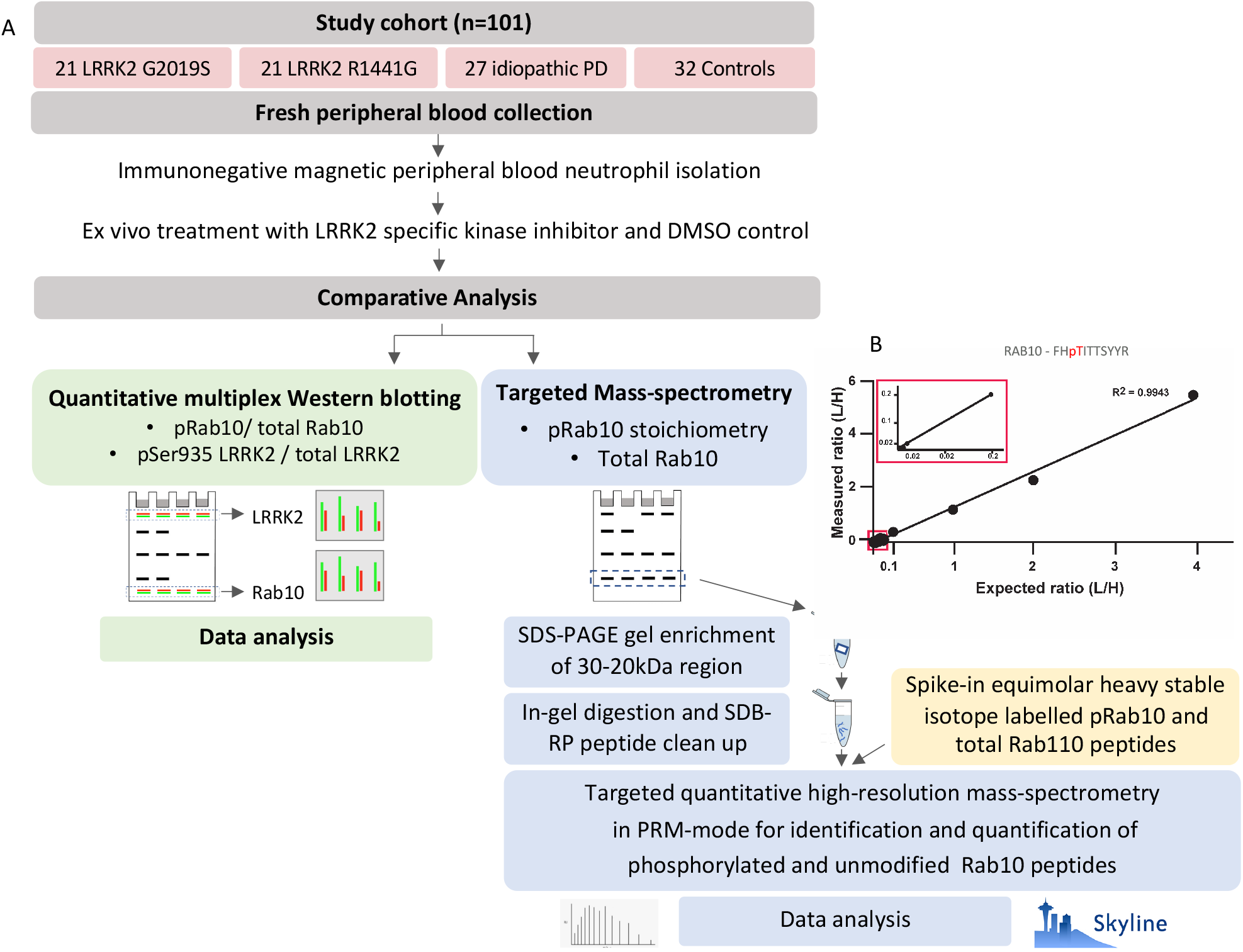
Experimental design and workflow for peripheral blood neutrophils. (A). 20 ml of fresh peripheral blood was collected from 101 participants with and without either PD or a pathogenic LRRK2 mutation – LRRK2 G2019S or LRRK2 R1441G – for peripheral blood neutrophil isolation by immunomagnetic negative isolation. Purified neutrophils were then split into two parts for *ex vivo* treatment with either the specific LRRK2 kinase inhibitor MLi-2 (200nM) or DMSO prior for 30 minutes prior to cell lysis and storage at −80 degrees Celsius. All sets of neutrophil lysates (MLi-2 and DMSO treated) were then subjected to quantitative multiplexed immunoblotting for Rab10^Thr73^ phosphorylation/total Rab10 protein ratio as well as targeted mass-spectrometry for Rab10^Thr73^ phosphorylation stoichiometry with spike in of heavy-labeled Rab10^Thr73^-phospho- and total peptide standards after enrichment SDS-PAGE followed by in-gel digestion followed by data analysis. Additionally, quantitative immunoblot analysis was performed for Serine 935 phosphorylation/ total LRRK2 protein as well as total Rab10 and total LRRK2 protein levels. **Limit of detection of targeted MS assay (B)**. Scatter plot depicting the Limit of detection and quantification of pRab10 in targeted PRM. 50 fmol of heavy pRab10 was mixed with a variable amount of light pRab10 ranging from 0.01, 0.1, 1, 10,50,100 and 200fmol that was spiked into 50ng HeLa. Light/Heavy ratio values were plotted to show the linear response of pRab10 (0.01 to 200fmol (R^2^=0.9943)). The zoom in rectangular box depicting the values of 0.01to 0.1fmol). n=3, error bars representing mean and SD.

### LRRK2 R1441G mutation carrier status significantly augments LRRK2 dependent Rab10^Thr73^ phosphorylation in neutrophils derived from PD manifesting and non-manifesting individuals

We analysed LRRK2 dependent phosphorylation of Rab10^Thr73^ in peripheral blood neutrophils from 42 LRRK2 mutation carriers in comparison to 32 controls as well as 27 iPD patients. Peripheral blod neutrophils were isolated from fresh blood by immunomagnetic negative selection as previously described(*19*). Prior to cell lysis neutrophils were split in half and treated with and without the LRRK2 kinase inhibitor MLi-2 to demonstrate that phosphorylation of Rab10 is mediated by LRRK2. When analysed by quantitative multiplex immunoblotting, we observed a striking 4.5-fold increase in LRRK2 dependent Rab10^Thr73^ phosphorylation in R1441G mutation carriers compared to controls, iPD and G2019S mutation carriers (Figure 2a). When segregated according to clinical disease status, Rab10^Thr73^ phosphorylation status was equally augmented in PD manifesting as well as non-manifesting individuals with R1441G positive mutation status (Figure 2a and Table 2). There was no statistically significant difference in Rab10^Thr73^ phosphorylation levels in G2019S mutation carriers irrespective of disease status and also not in iPD when compared to controls or with each each other. Total Rab10 levels were remarkable consistent amongst participants and did not differ between the groups (Figure 2b). A representative result of one technical replicate with duplicate loading for both DMSO and MLi-2 treated samples of the immunoblot analysis of all 101 participants is shown in Supplementary Figure S1. This also clearly shows the LRRK2 dependency of the Rab10^Thr73^ phosphorylation signal with significant reduction in the LRRK2 kinase inhibitor treated samples of all participants when compared to DMSO.

**Figure 2.**
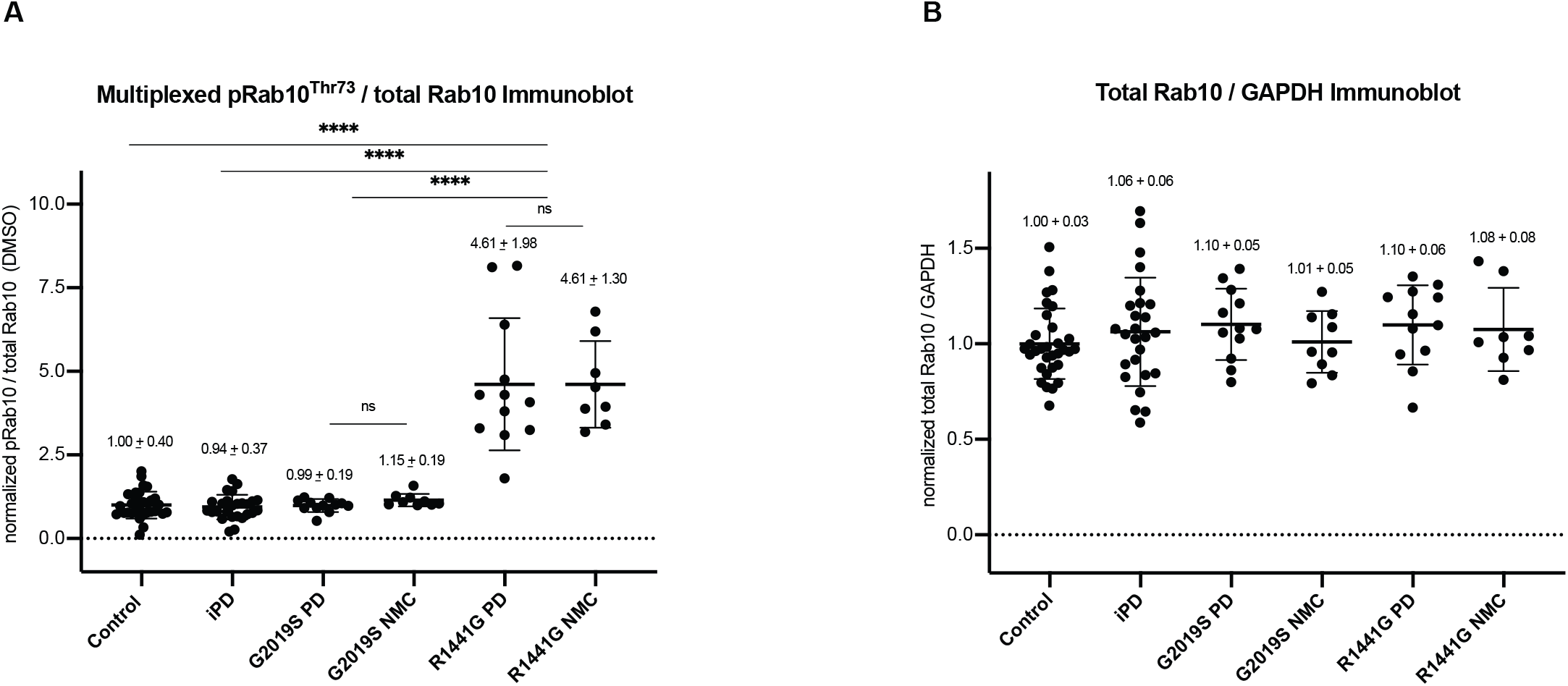
LRRK2 R1441G mutation carrier status significantly augments LRRK2 dependent Rab10Thr73 phosphorylation in neutrophils derived from PD manifesting and non-manifesting individuals. (A) Grouped analysis of Rab10^Thr73^ phosphorylation levels obtained by multiplexed immunoblotting with *MJFF*-*pRAB10* monoclonal antibodies against the LRRK2 phosphorylated Rab10 phosphoepitope Threonine 73 normalized to the total Rab10 protein levels of DMSO treated neutrophil lysates and to the average of the respective controls. Bars depict group means and standard deviation (SD). Quantifications were based on the average value of at least 2 independent immunoblot runs. The LRRK2 mutation carrier groups were further broken down by clinical disease status – either PD-manifesting (PD) or non-manifesting carriers (NMC). Differences between groups were calculated by one-way ANOVA followed by multiple comparisons where the mean of each column was compared against the mean of all other groups. **** p<0.000. (B) Total Rab10 protein levels normalized against the housekeeping protein GAPDH (mean and SD).

### Positive correlation between Western blotting and targeted mass-spectrometry assays for LRRK2 dependent Rab10 phosphorylation

In parallel to quantitive immunoblotting for Rab10^Thr73^ phosphorylation, the neutrophil samples were subjected to a ultra-sensitive targeted mass-spectrometry (MS) based assay for analysing Rab10^Thr73^ phosphorylation stoichiometry using synthetic stable isotope-labelled (SIL) tryptic Rab 10 peptides around the Thr73 epitope in its phosphorylated as well as unphosphorylated form as internal standards (*18*) (Figure 1). This analysis allowed us to addess reproducibility of our immunoblotting results by an independent method, correletion between the 2 assays and importantly explore whether a potentially more sensitive analysis by mass-spectrometry could yield a statistically significant difference in the G2019S mutation carrier group or iPD patients when compared to controls.

We measured Rab10^Thr73^ phosphorylation occupancies in all MLi-2 treated and untreated (DMSO) neutrophil samples from all 101 participants (Figure 3a). Consistent with immunoblotting data, the average Rab10^Thr73^ occupancies in the control group were 1.52% whereas they increased over 4-fold to 6.60% in the R1441G mutation carrier group (p<0.0001,one-way Anova). Here again, the MS assay confirmed that there was no significant difference between control group occupancies in comparison to the iPD (1.61%) and G2019S mutation carrier groups (1.90%) (Table 2). Furthermore, when mutation carriers were separated by clinical PD status and analysed by unpaired t-testing there was also no difference between R1441G PD manifesting and non-manifesting individuals (p=0.23) or between G2019S PD manifesting and non-manifesting individuals (p=0.33) (Figure 3a and Table 2b). Figure 3a also shows the respective Rab10^Thr73^ phosphorylation occupancies in the LRRK2 kinase inhibitor treated samples (MLi-2) which was significantly reduced when compared to the untreated (DMSO) samples for each group (p<0.0001, Table 2b) and therefore clearly demonstrating that the phosphorylation of Rab10 is mediated by the LRRK2 kinase. As with the analysis by Western blotting, mass-spectrometry analysis for total Rab10 levels did not significantly differ between groups (Figure 3b).

**Figure 3.**
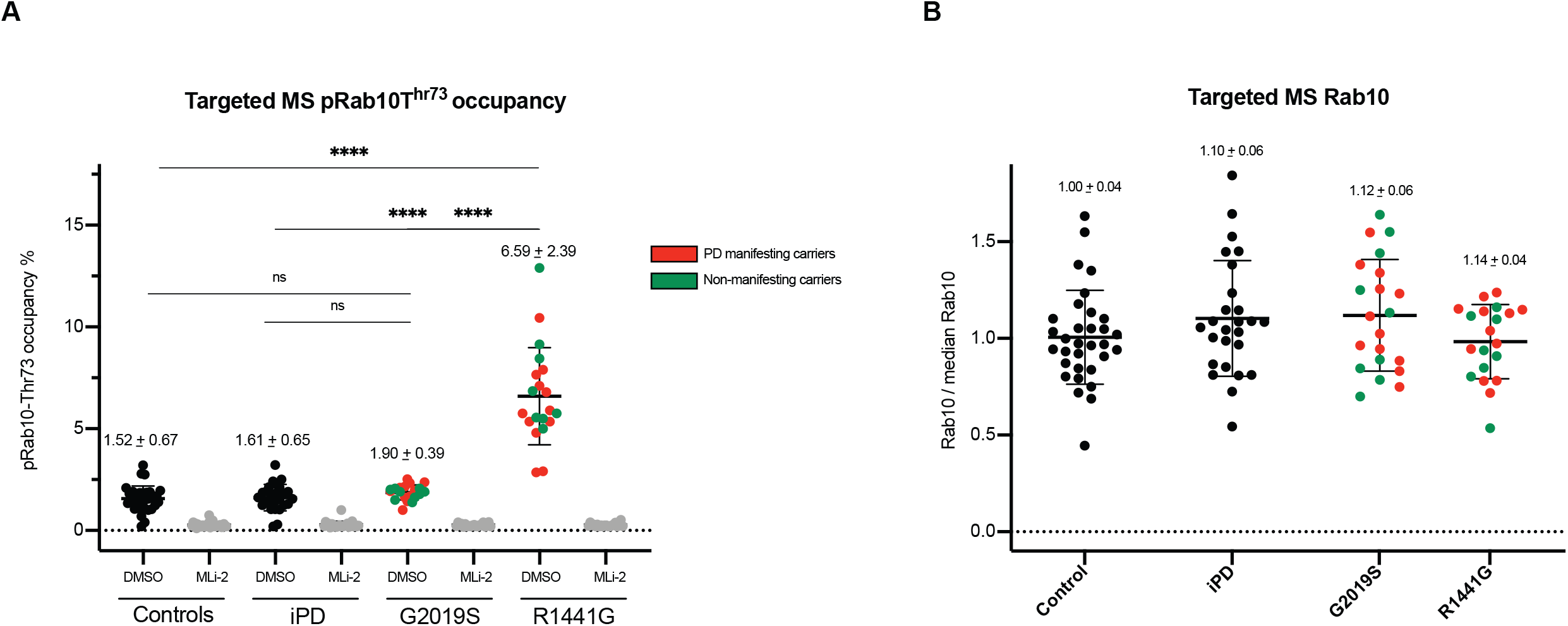
Significantly elevated Rab10-pThr73 phosphorylation stoichiometry in neutrophils derived from R1441G mutation carriers with and without PD. (A) Quantification of Rab10-pThr73 occupancy (%) in DMSO and MLi-2 treated neutrophil lysates derived from controls, iPD and LRRK2 mutation carriers of either the G2019S or R1441G mutation. PD or non-manifesting carriers (NMC) status indicated in DMSO samples by colour. One-way ANOVA with multiple comparisons was applied with the mean of each column being compared with the mean of the control group. Rab10^Thr73^ phosphorylation occupancy is presented as means1.±1.SD. **** p<0.000. There was no statistically significant difference between manifesting and non-manifetsing mutation carriers for both R1441G and G2019S mutation (see also table 2a). (B) Peak areas of endogenous total Rab10 (two peptides) were summed and normalized to median intensity displayed with mean and SD. Proteomic analysis of total Rab10 peptide levels did not differ between groups.

While the targeted Rab10^Thr73^ phosphorylation mass-spectrometry analysis of our neutrophil samples independently confirmed that neutrophils from R1441G, but not G2019S mutation carriers or patients with iPD have significantly elevated LRRK2 dependent Rab10 phosphorylation levels, we were interested in directly comparing the results obtained by the 2 methods. We therefore aligned the MS and WB data per individual (Figure 4a and 4b) and found good correlation between the 2 methods (R squared = 0.78, p<0.0001)(Figure 4c). Representative results are shown in Figure 5 where duplicate DMSO and MLi-2 treated neutrophil samples from the same 10 individuals including 5 controls and 5 R1441G mutation carriers were analysed for pRab10 levels by quantitative Western blotting (Figure 5a) and targeted pRab10 mass-spectrometry (Figure 5b). The data also clearly shows that the Rab10 phosphorylation signal is significantly reduced to almost undetectable levels when neutrophils were treated with the specific LRRK2 kinase inhibitor MLi-2 confirming the specificity of the pRab10 signal as a substrate of the LRRK2 kinase in this cell type.

**Figure 4.**
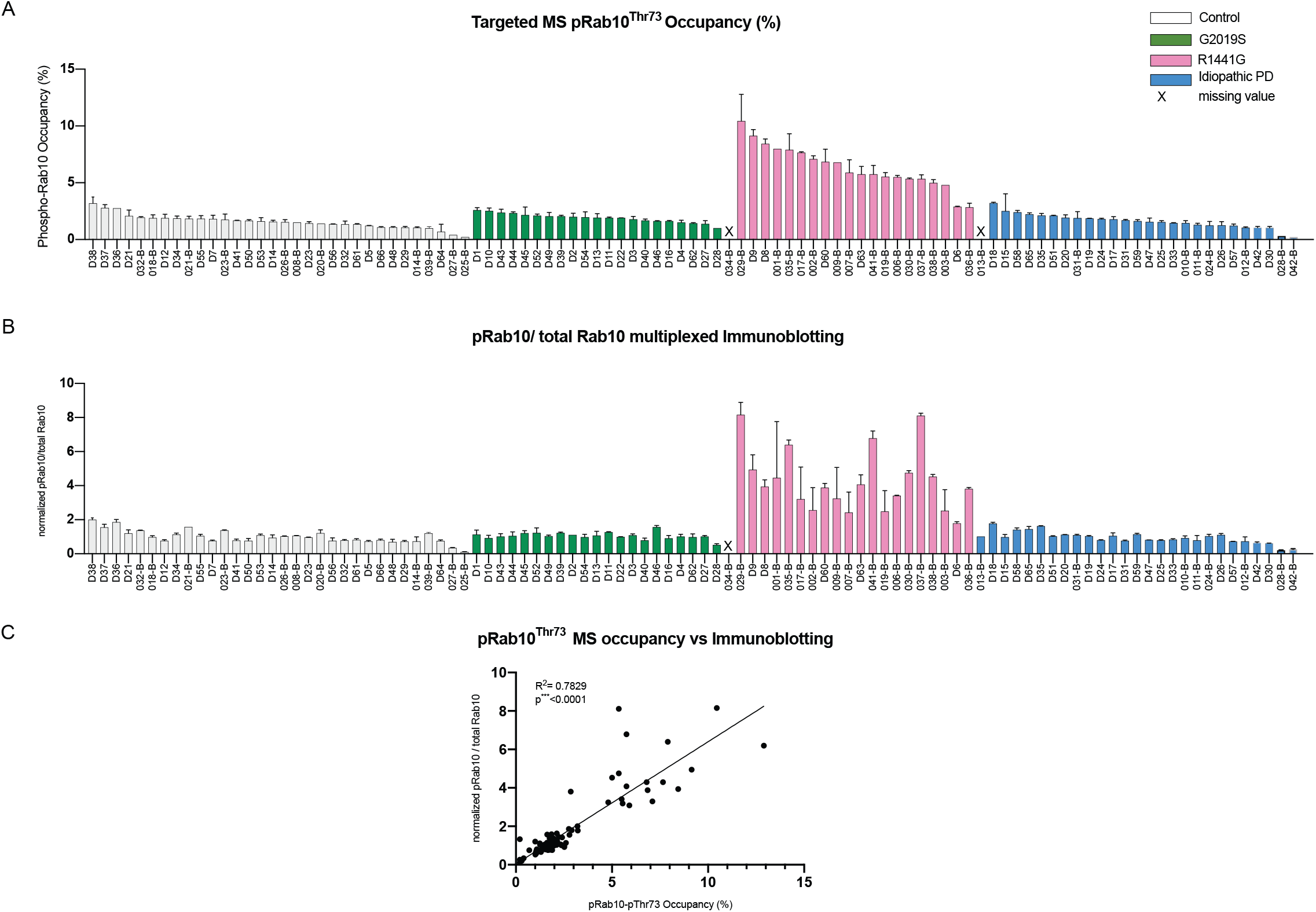
Good correlation between quantitative multiplexed phosphorylated Rab10Thr73/total Rab10 immunoblotting and targeted Rab10^Thr73^ occupancy assays in peripheral blood neutrophils. (A) Rab10^Thr73^ phosphorylation stoichiometries in DMSO treated neutrophils per individual arranged in descending order by group (B) and aligned quantitative immunoblotting analysis of pRab10/total Rab10 ratios (B) of 101 donors were plotted as mean with SD for all 101 participants. (C) Pearson correlation between immunoblotting for Rab10^Thr73^ phosphorylation/total Rab10 protein levels and targeted Rab10^Thr73^ occupancies. Blotted are mean values of 2 independent experiments for each method (R^2^=0.78, p<0.0001).

**Figure 5.**
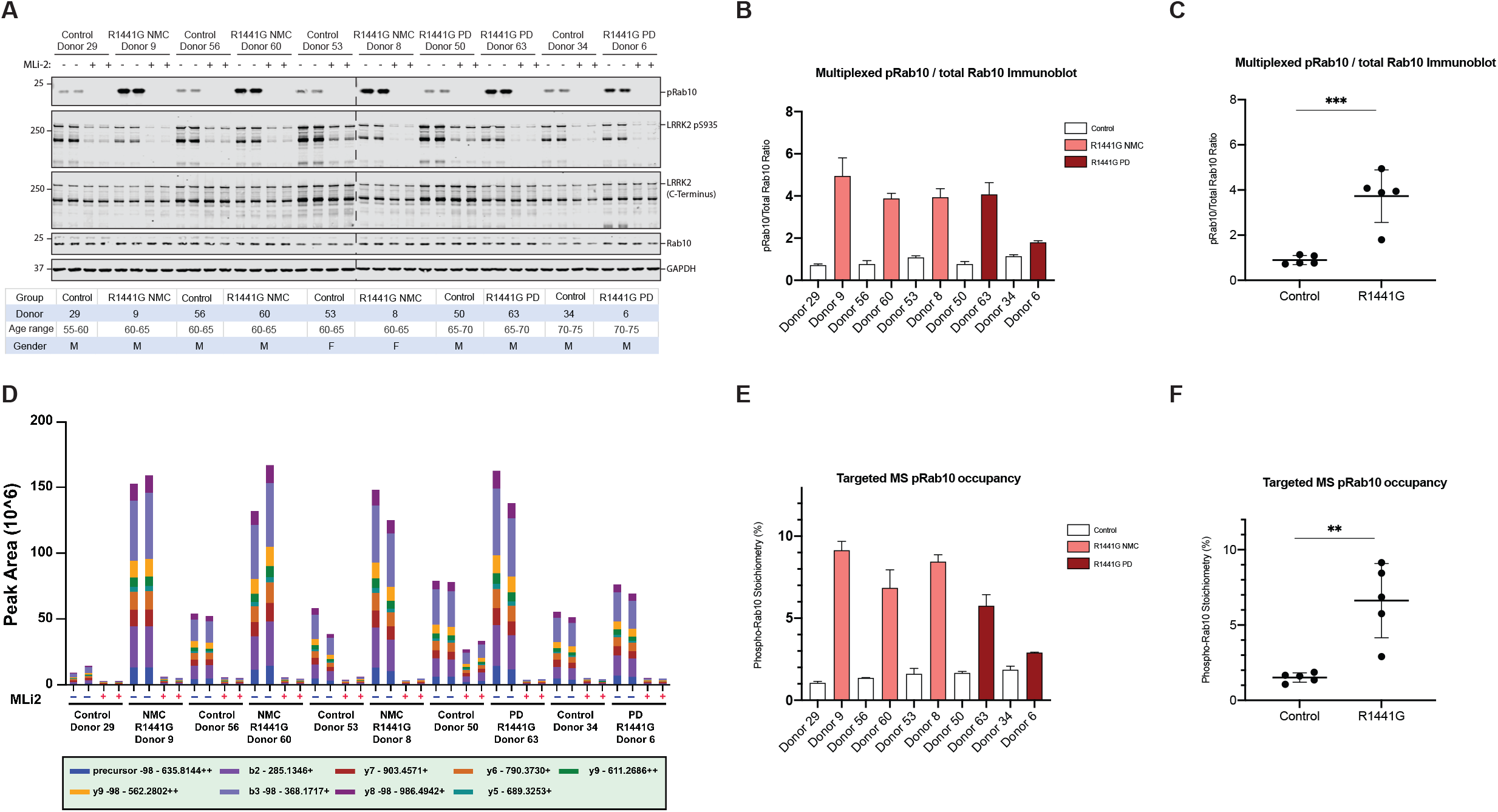
Representative results for 5 controls and 5 R1441G mutation carriers. (A) Immunoblotting of neutrophils isolated from 5 R1441G LRRK2 mutation carriers with PD (n=2) and non-manifesting carrier status (n=3) as well as 5 controls with duplicate loading of 10 µg of DMSO and MLi-2 (200 nM MLi-2, 30 min) treated whole cell extracts using antibodies against total LRRK2, pSer935 LRRK2, total Rab10, MJFF-pRAB10 (pThr73) and GAPDH. (B) Quantification of immunoblots of 2 independent experiments by analyzing phospho-Thr73 Rab10/total Rab10 ratio of DMSO treated samples (B) per individual and (C) per group with group difference calculated by unpaired t-test (***P=0.0007). (D) The respective summed intensities of fragment ion transitions are represented with different colours as shown in the bottom panel of the graph. (E) Relative endogenous pRab10 peak areas of 2 independent analysis for pRab10^Thr73^ occupancy depicted as a bar graph for DMSO and MLi-2 treated samples of all 10 participants and (F) per group with group difference calculated by unpaired t-test (**P=0.0014).

### LRRK2 total protein and Serine 935 phosphorylation levels in peripheral blood neutrophils

In addition to phosphorylated and total Rab10 levels, we analysed Serine 935 phosphorylation and total LRRK2 levels by quantitative Western blotting in all peripheral blood neutrophil samples. We observed a small, but statistically significant reduction in Serine 935 phosphorylation in R1441G mutation carriers when compared to iPD (p=0.045), but no difference when compared to G2019S mutation carriers or controls (Figure 6a). This is consistent with previous work that has revealed that the LRRK2 R1441G mutation reduces Ser935 phosphorylation(*13, 16*). When Serine 935 phosphorylation levels were compared between R1441G mutation carriers with and without PD, there was no difference (p=0.99). We did not find a statistically significant reduction in G2019S mutation carriers irrespective of disease status when compared to iPD or in fact any of the other groups. LRRK2 total protein levels did not significantly differ between the R1441G mutation carriers, G2019S mutation carriers, iPD and controls (Figure 6b).

**Figure 6.**
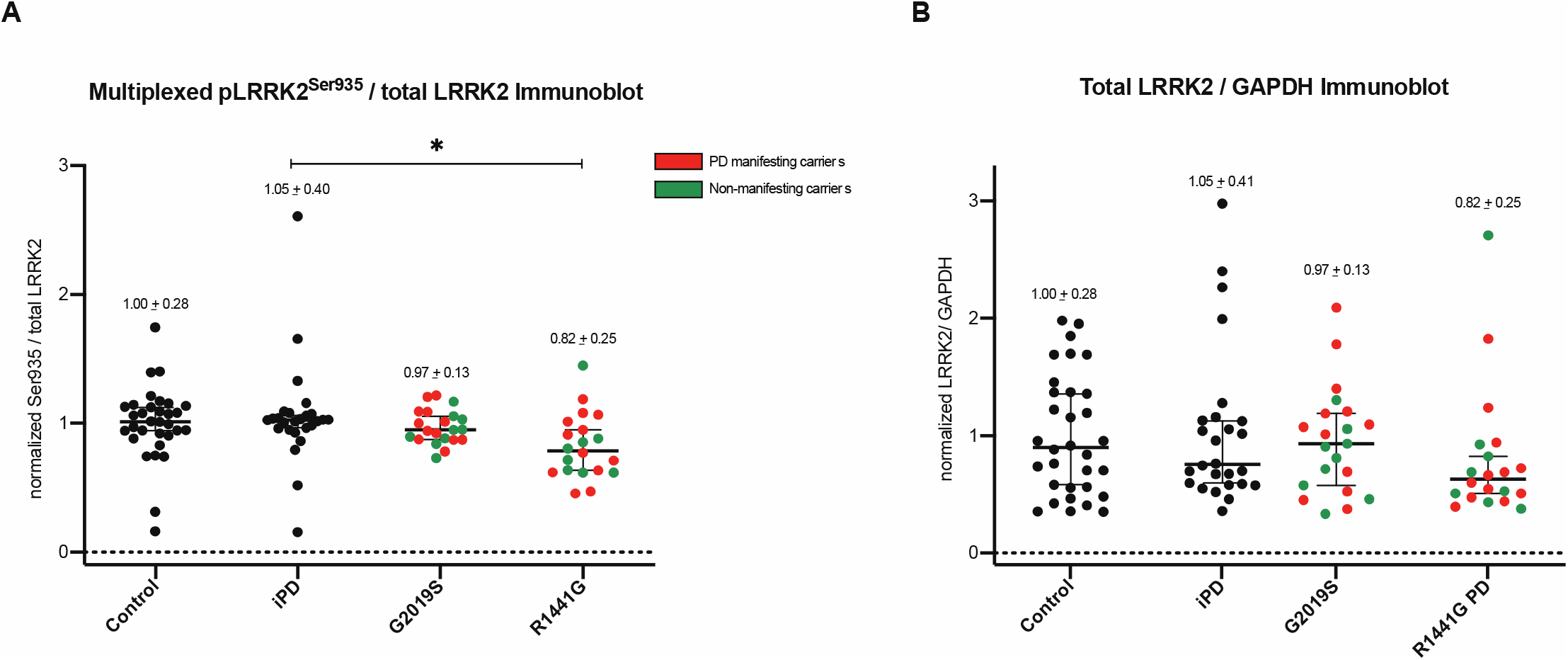
pSerine935 and total LRRK2 levels in peripheral blood neutrophils. Normalized multiplexed Serine935 / total LRRK2 protein ratio (A) and normalized total LRRK2 / GAPDH protein levels (B) of DMSO treated neutrophil lysates per group. Bars depict mean values with SD. Quantifications were based on the average value of 2 independent immunoblot runs. Differences between groups were calculated by one-way ANOVA followed by multiple comparisons where the mean of each column was compared against the mean of the other groups. There was a small, but statistically significant difference between the R1441G mutation carrier and iPD groups *p=0.0454 for Serine 935/total LRRK2 while total LRRK2 / GAPDH ratio remained largely unchanged between groups. When segregated by disease status, there was no significant difference between PD manifesting and non-manifesting mutation carriers for G2019S and R1441G.

### Rab10^Thr73^ phosphorylation in human post-mortem brain samples

PD is considered a condition of the central nervous system and an important question is how the analysis of LRRK2 dependent Rab10 ^Thr73^ phosphorylation in human peripheral blood neutrophils compares to the analysis of Rab10 ^Thr73^ phosphorylation and LRRK2 levels in the brain. In order to address this, we obtained matched frontal and occipital cortex samples from 10 individuals including 3 controls, 4 G2019S mutation carriers with PD and 3 iPD from the brain bank at the Columbia University Medical Center in New York. Additionally, we were able to access 20 frontal cortex samples including 7 controls, 7 iPD, 5 G2019S and 1 R1441H mutation carriers with PD from the IDIBAPS Biobank at the Hospital Clinic in Barcelona, Spain. Demographic and clinical information is provided in Supplementary table 3. We used multiplexed quantitative immunoblotting to assess Rab10^Thr73^ phosphorylation against total Rab10 levels and Serine 935 phosphorylation against total LRRK2. Total protein levels for Rab10 and LRRK2 were measured by normalization against a housekeeping protein. With regards to Rab10^Thr73^phosphoryation, we observed high variability amongst samples from low to almost undetectable levels in the majority of samples and very high levels in some irrespective of group or LRRK2 genotype; in fact, the highest levels were observed amongst the control samples. Also, the R1441H genotype, which is as R1441G another hotspot mutation at the R1441 site and known to significantly augment LRRK2 dependent Rab10 phosphorylation in the heterologous overexpression system (unpublished results), did not result in significantly elevated Rab10 Rab10^Thr73^ phosphorylation levels. While the variances were higher for the control samples (standard deviation = 8.233 vs. 2.495 (LRRK2 mutation carriers) and 2.640 (iPD)), there were no significant differences between the control, LRRK2 mutation carrier and iPD groups. Total Rab10 levels displayed less variability amongst frontal cortex samples and did not significantly differ between the groups. There was also a high degree of variability in total LRRK2 and Serine 935 phosphorylation levels amongst samples, but overall no significant difference between the LRRK2 mutation carrier, iPD and control groups (Supplemental figure S3). When comparing frontal and occipital lobe samples where matched pairs were available, Rab10^Thr73^ phosphoryation tended to be lower in the occipital lobe than the frontal lobe (with the exception of one G2019S mutation carrier (Col-3) where the levels in the occipital lobe were higher) and total Rab10 levels were always lower in the occipital lobe samples (p=0.0016). For Serine 935 phosphorylation, there was no significant difference between matched frontal and occipital lobe samples (paired t-test, p=0.3235), while total LRRK2 levels were significantly reduced in the occipital lobe when compared to matched frontal lobe samples (p=0.001). Additionally, we subjected the brain lysates to immunoblotting for the PPM1H phosphatase that counteracts LRRK2 signalling by selectively dephosphorylating Rab proteins including Rab10(*27*), but did not observe significant significant changes amongst samples or groups except for reduced levels in sample Col-3.

## Discussion

This is to date the largest study of LRRK2 dependent phosphorylation of its endogenous substrate Rab10^Thr73^ in peripheral blood neutrophils in LRRK2 mutation carriers, controls and individuals with iPD. Our results include 3 key observations: 1) We show for the first time that LRRK2 dependent Rab10^Thr73^ phosphorylation as a surrogate marker for LRRK2 kinase activation status is significantly elevated in carriers of a specific pathogenic variant in the LRRK2 gene, namely the R1441G mutation in the ROC-COR GTPase domain. 2) This significant increase in Rab10^Thr73^ phosphorylation in R1441G mutation carriers is irrespective of clinical disease status and observed in non-manifesting carriers as well as those diagnosed with PD. 3) We have not observed a statistically significant increase in LRRKK2 dependent Rab10^Thr73^ phosphorylation in carriers of the common G2019S mutation deploying 2 independent assays. Additionally, we show that both Serine 935 as well as Rab10^Thr73^ phosphorylation serve as biomarkers for LRRK2 kinase inhibition when using kinase inhibitors such as MLi-2. Our explorative analysis of markers of the LRRK2 kinase pathway in human autopsy brain samples highlights important aspects and indeed problems with regards to feasibility, suitability and differences across different biomatrices.

Utilizing our facile and robust multiplex immunoblotting assay using sensitive phosphospecific antibodies against the Rab10 Threonine 73 site that is phosphorylated by the LRRK2 kinase, a statistically significant increase of more than 4-fold in Rab10^Thr73^ phosphorylation levels was seen in LRRK2 R1441G mutation carriers when compared to iPD, G2019S mutation carriers or controls. This increase was irrespective of disease status and there was no significant difference between R1441G mutation carriers with or without PD manifestation. This suggests that the effect on Rab10^Thr73^ phosphorylation levels in circulating neutrophils is mainly driven by the underlying mutation itself rather than by the accompanying Parkinson’s disease process. In future work it would be interesting to monitor larger numbers of particpants and over time to better assess the utility of LRRK2 dependent Rab10^Thr73^ phosphorylation as a marker of LRRK2 driven disease conversion or disease state. It would also be interesting to explore additional biomatricies.

Heterologous expression systems such as HEK293 overexpression of LRRK2 variants and genetic animal models with knock-in of LRRK2 mutations in the homozygous state have shown that the LRRK2 Roc and Cor domain mutations – including R1441G/H that suppress GTPases activity and promote GTP binding-mediate a 3-4-fold increase in LRRK2 kinase activity(*14, 15*) while the LRRK2 kinase domain mutations such as G2019S increase LRRK2 kinase activity only modestly, typically under 2-fold (reviewed in(*13*)) by domain disruption. In our experiments, we did not observe a statistically significant increase in Rab10^Thr73^ phosphorylation level in 21 G2019S mutation carriers despite promising results from a previous much smaller study of Rab10^Thr73^ phosphorylation stoichiometry in neutrophils from 4 G2019S mutation carriers in comparison to 4 non-mutation carriers(*18*). While there was good correlation between the 2 assays which allowed for independent confirmation of our results, the potentially more sensitive mass-spectrome assay did not yield any additional information with regards to LRRK2 dependent Rab10^Thr73^ phosphorylation levels. Given that the LRRK2 G2019S mutation induces an under 2-fold enhancement of Rab10 phosphorylation in homozygosity, one would expect a relative reduction to <1.5-fold in heterozygosity. We therefore conclude that the effect of the G2019S mutation in heterozygosity as with the participants in our study coupled with the biological variation in between human samples is too small to accurately yield a significant difference by quantitative immunoblotting or even more sensitive MS methodologies.

A recent study found a statistically significant decrease in LRRK2 Serine 935 phosphorylation in human peripheral blood mononuclear cells (PBMCs) derived from LRRK2 G2019S mutation carriers with PD but not G2019S NMC when compared to iPD (*30*). In this study we have not observed a reduction in Serine 935 phosphorylation levels in G2019 mutation carriers with PD in comparison to iPD or in fact to any of the other groups. However, the differences in biomatrix – PBMCs vs neutrophils – and assay methodology – digital immunoassay vs quantitative Western blotting – may account for this discrepancy. For example, neutrophils represent a homogenous cell population with high LRRK2 expression, but their intrinsically very active serine proteases appear to affect disproportionately high molecular weight species such as LRRK2 (288kDa) with resulting partial LRRK2 degradation(*20*). This particular problem could be addressed by using peripheral blood monocytes for LRRK2 pathway analysis as described by us previously(*26*). It would therefore be interesting to expand and compare Serine 935 phosphorylation across different peripheral blood cell populations including monocytes and additional patient cohorts. What we did observe was a reduction in Serine 935 phosphorylation in R1441G mutation carriers when compared to iPD, but not when segregated by clinical status or in comparison to G2019S mutation carriers or controls, which as mentioned above is consistent with previous studies (*13, 31*).

Rab10^Thr73^ phosphorylation has previously been shown to be elevated in substantia nigra of patients with iPD when compared to controls (*32*). While we were interested in extending our analysis to postmortem brain samples of LRRK2 mutation carriers and iPD in comparison to controls and assess its correlation with peripheral blood neutrophils, we were unable to obtain substania nigra samples, but only frontal and additionally occipital cortex in some. While total Rab10 protein levels were relatively stable amongst all samples, the Rab10^Thr73^ phosphorylation signal was not or hardly detectable in the majority of cases and otherwise variable irrespective of genotype and clinical disease status. In particular, we did not observe significant Rab10^Thr73^ hyperphosphorylation in the one LRRK2 R1441H mutation carrier sample which biochemically should have the same augmenting effect on LRRK2 dependent Rab10^Thr73^ phosphorylation as the R1441G mutation(*14*). While the LRRK2 total protein and Serine 935 phosphorylation signals could be detected there was again a large degree of disparity in between samples. Overall, the main finding was that there was high variability amongst individual samples in all parameters that we analysed. LRRK2 pathway components including Rab10 phosphorylation have been previously analyzed in another study of human brain cingulate cortex samples from 23 controls and 28 idiopathic Parkinson’s patients(*5*). This study also revealed variable levels pRab10 and LRRK2 between different subjects. While it would be worthwhile to expand the analysis of Rab10^Thr73^ phosphorylation in autopsy brain material to a larger number of cases, in particular of carriers of R1441 hotspot or the VPS35 D620 mutations, there are important factors to consider such as post-mortem interval that may have a different effect on posttranslational modifications when compared to the unmodified protein or indeed higher molecular weight species such as LRRK2(*33*). Another concern above and beyond the regional variation of brain tissue is its heterogeneity in terms of cellular composition which is not taken into account in the assays deployed in this study. Nevertheless, there is ample evidence derived from genetic mouse models with knock-in of either the LRRK2 R1441C or VPS35 D620N mutations, that the increase in LRRK2 dependent Rab10^Thr73^ phosphorylation levels in peripheral organs such as spleen, lung or kidneys as well as cultured fibroblasts corresponds to equally augmented LRRK2 dependent Rab10^Thr73^ phosphorylation in brain(*5, 26*).

In conclusion our findings add compelling evidence that interrogating LRRK2 dependent Rab10 phosphorylation in human peripheral blood neutrophils is a robust and facile surrogate marker for LRRK2 kinase activation status and will identify individuals with an above 2-fold increase in LRRK2 kinase activity. Further work is required to develop more sensitive assays to detect the impact of mutations with more modest effect on LRRK2 kinase activity such as the common LRRK2 G2019S mutation and other LRRK2 variants. It may be necessary to better explore the impact that the G2019S mutation has on downstream biology and use this information to develop additional biomarkers. Neverthless, we envision that our LRRK2 dependent Rab10 phosphorylation assays will be used alongside other markers of the LRRK2 and PD associated pathways across different biomatrices and importantly longitudinally to possibly delineate integrated markers for disease conversion and PD progression. In combination with genetics our assay can serve as a functional marker to identify novel mutations and genes that exert their effect via activation of the LRRK2 kinase signalling network.

## Data Availability

All data is available

The Authors declare that there are no competing interests associated with the manuscript.

## Funding

The work was supported by funding from the Michael J. Fox Foundation for Parkinson’s Research, small grant funding from Parkinson’s UK, and Medical Research Councile (Dario Alessi). Esther Sammler was supported by the Tayside medical Science Centre (TASC) and a Scottish Senior Clinical Academic Fellowship.

Ethics approval was granted by the respective Institutional Review Boards at the University Hospital in Barcelona and Biodonostia Health Research Institute in San Sebastian, Spain and the University of Columbia in New York, US. Tissue and lysates were stored and used at the University of Dundee in line with regulations from the Tayside Tissue bank.

**Supplementary Table S1**. List of materials and reagents.

**Supplementary Table S2. Detailed demographic and clinical characteristics of peripheral blood neutrophil donors**. Including group (LRRK2-PD, LRRK2-non-manifesting carrier (NMC), idiopathic PD and control), study and original site ID, specific mutation that participant carriers, age range, disease duration, age at PD diagnosis, *LEDD* is L-dopa equivalent daily dosage, *UPDRS* is Unified Parkinson’s Disease Rating Scale, part III (motor symptoms) and part IV (motor complications), participant’s location site (either Barcelona or San Sebastian).

**Supplementary figure S1. Representative Immunoblot analysis of peripheral blood neutrophil samples**. Neutrophils isolated from fresh peripheral blood were treated with either DMSO vehicle control or the specific LRRK2 kinase inhibitor MLi-2 at a concentration of 200nM for 30 minutes prior to cell lysis. 10 µg of whole cell extracts were then loaded in duplicates and subjected to quantitative immunoblot analysis with the indicated antibodies and the membranes developed using the Odyssey CLx scan Western Blot imaging system. pRab10 and total Rab10 protein as well as Serine 935 and total LRRK2 antibodies were multiplexed and the same internal standard was run on every gel to compare samples run on different gels (not shown). Similar results were obtained in two independent immunoblot experiments of the same extracts. (A) Representative immunoblot for neutrophils derived from the 66 participants from Barcelona (ID 1-66). (B) Representative immunoblot for neutrophils derived from the 35 participants from San Sebastian (ID 67-101).

**Supplementary Table S3. Demographic characteristics of postmortem brain samples**. Demographic data for the brain tissue cases used in this study are shown including sample site (either Columbia or Barcelona), group including LRRK2 associated PD, PD or control, LRRK2 genotype, brain region, gender, age range, cold postmortem interval (PMI) and * frozen PMI where cold PMI was not available as well as histopathological assessment.

**Supplementary figure S2: Representative Immunoblot analysis in human brain extracts**. Snap frozen autopsy samples were obtained from matched frontal and occipital cortex samples of 10 individuals including 3 controls, 4 G2019S mutation carriers with PD and 3 iPD from the brain bank at the Columbia University Medical Center in New York, USA and additionally 20 frontal cortex samples from 7 controls, 7 iPD, 5 G2019S and 1 R1441H mutation carriers with PD from the IDIBAPS Biobank at the Hospital Clinic in Barcelona, Spain. Whole tissue lysates from each of these were generated and duplicate loading of 20 µg subjected to immunoblot analysis using the indicated antibodies. pRab10 and total Rab10 protein as well as Serine 935 and total LRRK2 antibodies were multiplexed and the same internal standard was run on every gel to compare samples run on different gels (not shown).

**Supplementary figure S3. Quantification of Rab10**^**Thr73**^ **phosphorylation levels in post-mortem brain samples from LRRK2 mutation carriers, iPD and controls**. Quantifications were based on the average value of 2 independent immunoblot experiments with duplicate sample loading. Quantification of phosphorylated Rab10^Thr73^/total Rab10 ratio and total Rab10 protein levels / GAPDH per sample with columns indicating means and SD (A), (C) and per group for the frontal cortex samples (B), (D), respectively. The same analysis was performed for Serine935 / total LRRK2 protein ratio and total LRRK2 / GAPDH protein levels per sample (E), (G) and per group for the frontal cortex samples (F), (H), respectively.The R1441H mutation carrier sample is marked in pink in the grouped analysis. Overall, there was no significant group difference.

