## Supplementary figures and images for "R1441G but not G201S mutation enhances LRRK2 mediated Rab10 phosphorylation in human peripheral blood neutrophils"

### Supplementary figure 1A

#### Slide 1
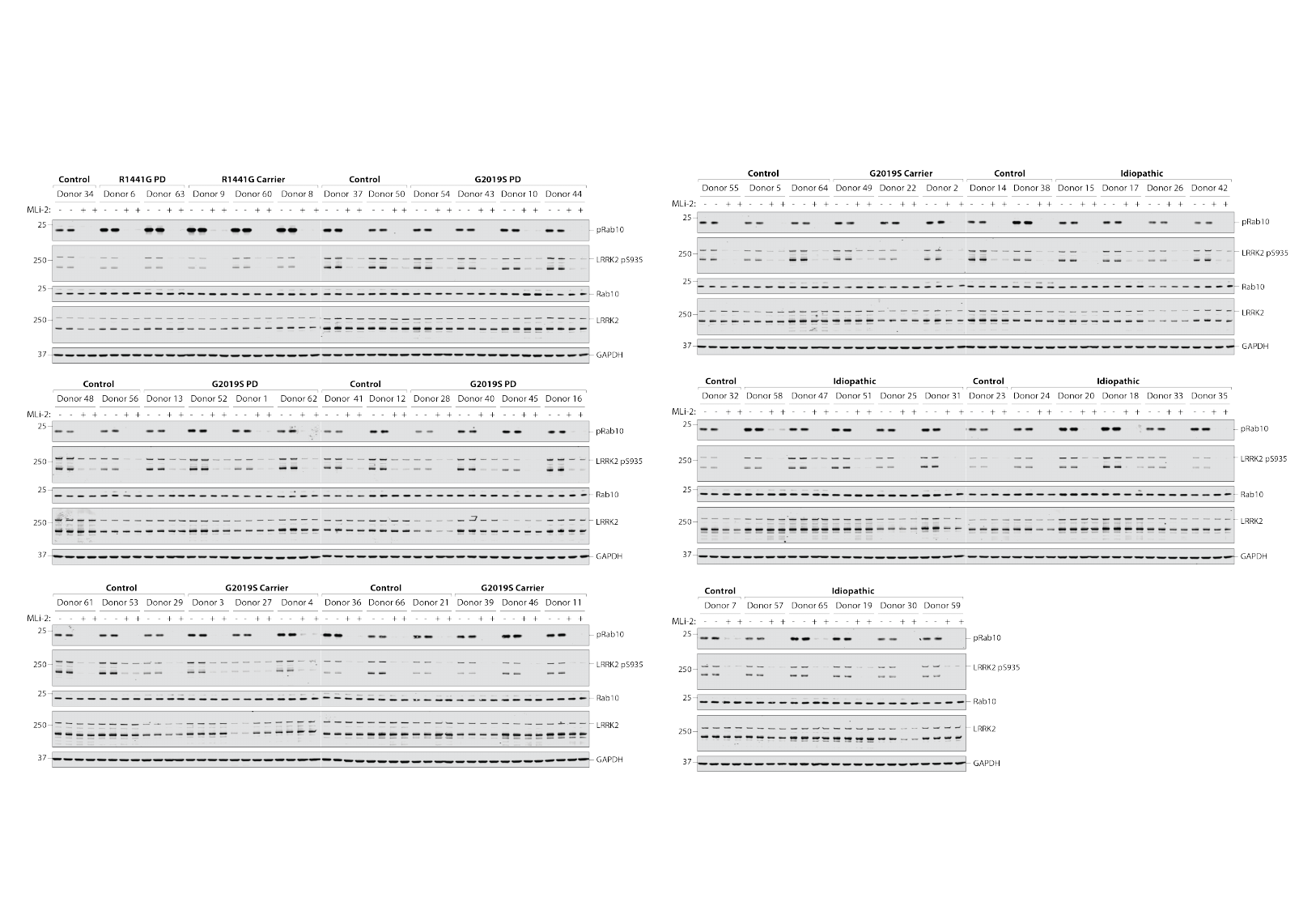

### Supplementary figure 2

#### Slide 1
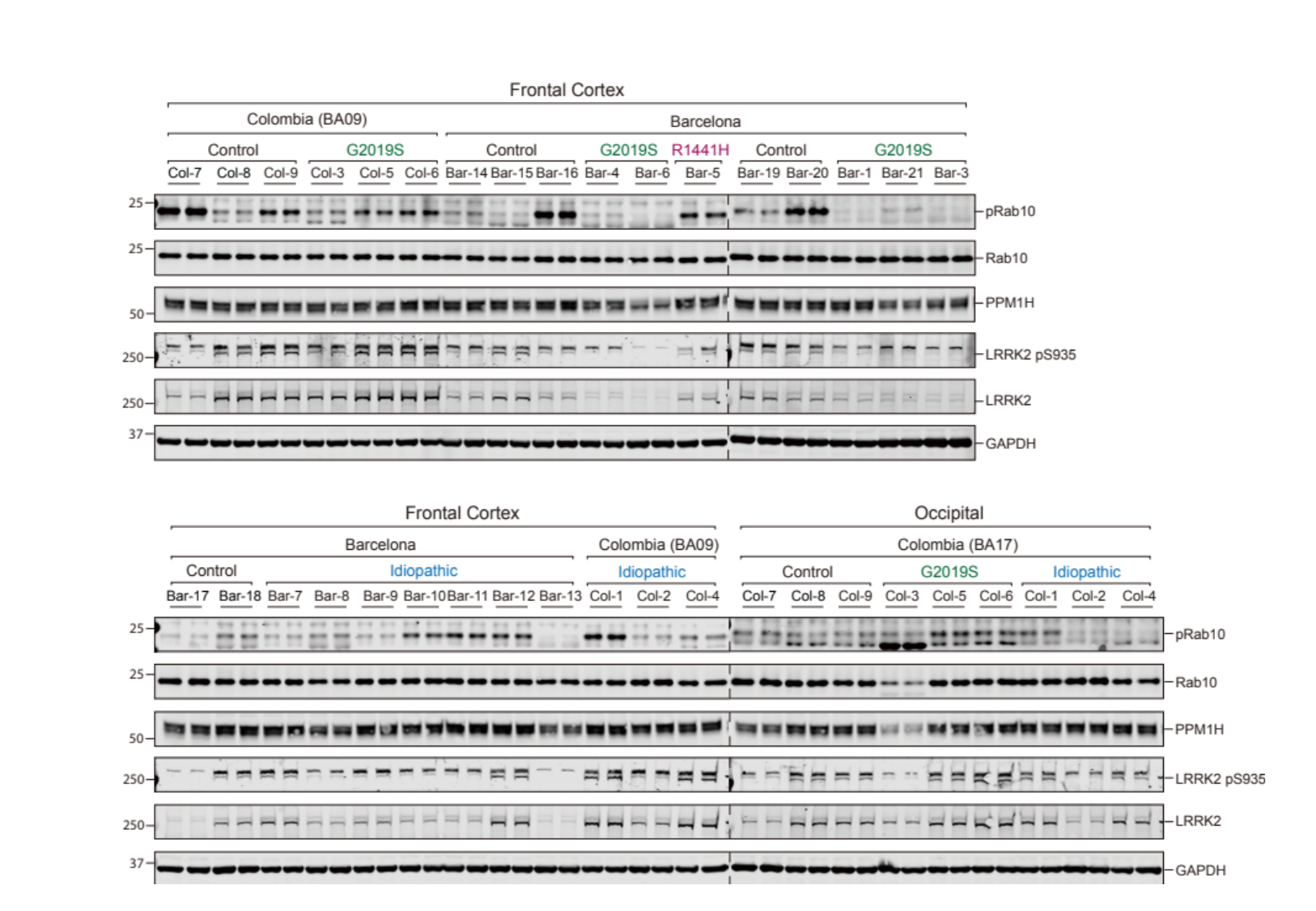

### Supplementary figure S1BC

#### Slide 1
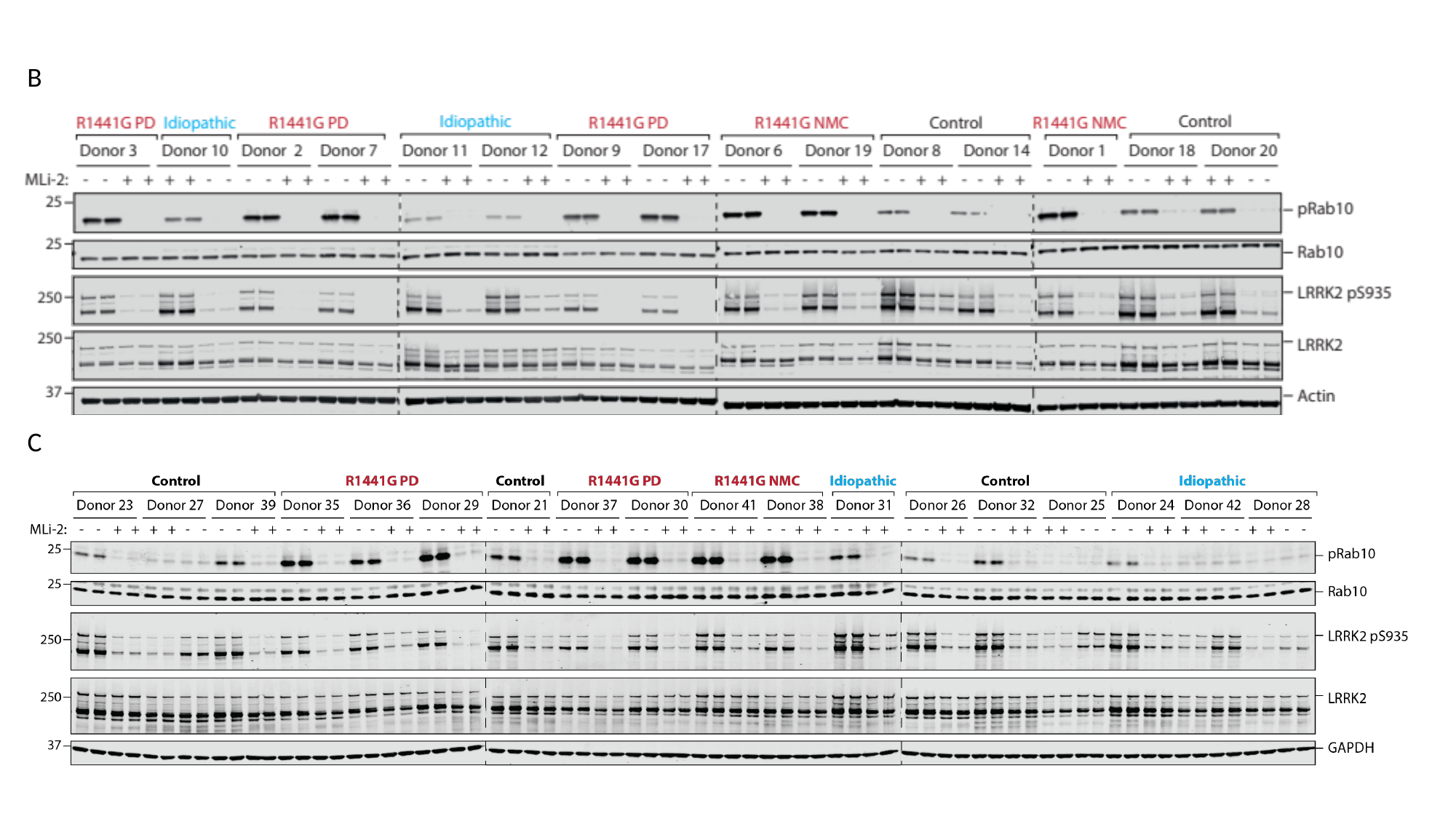

B
C

### Supplementary figure S3

#### Slide 1
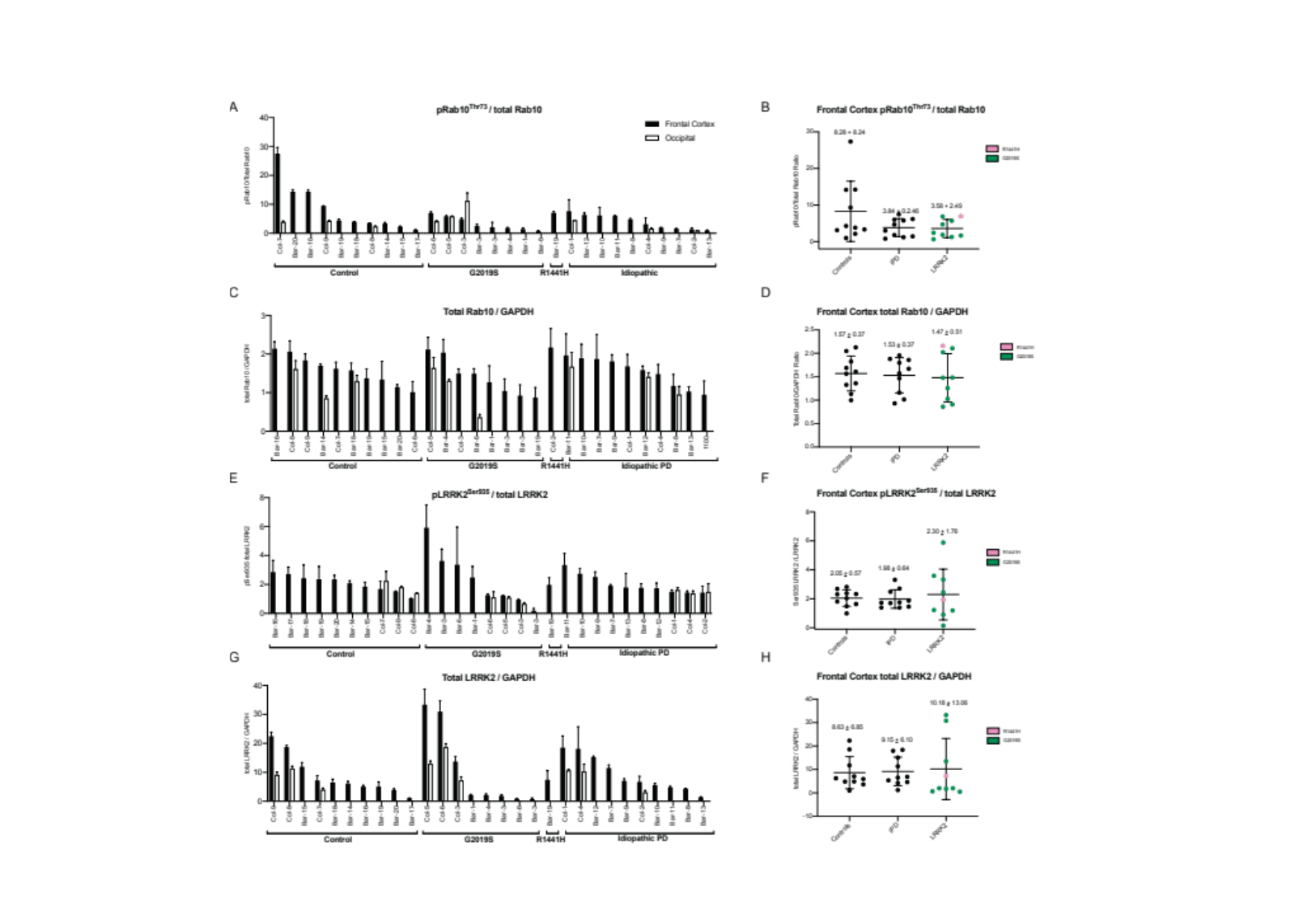

### Supplementary table S1

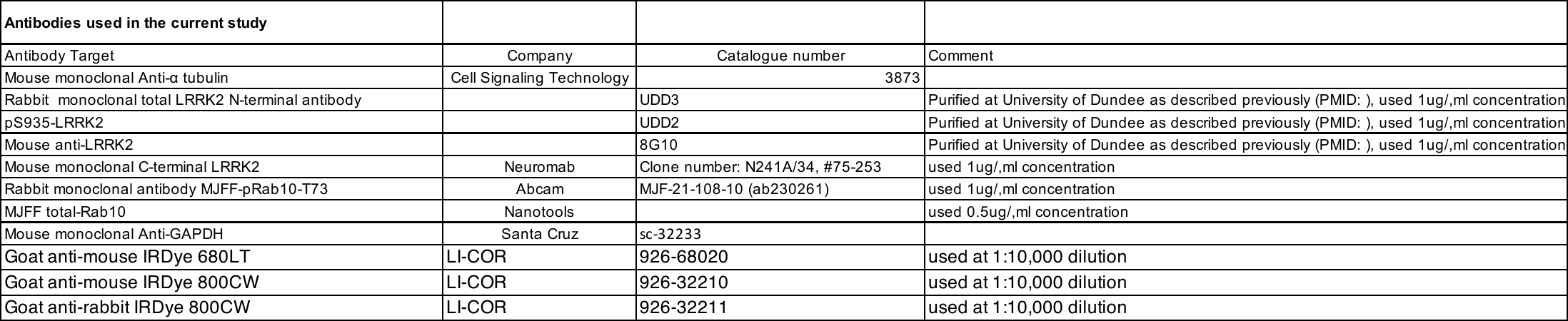

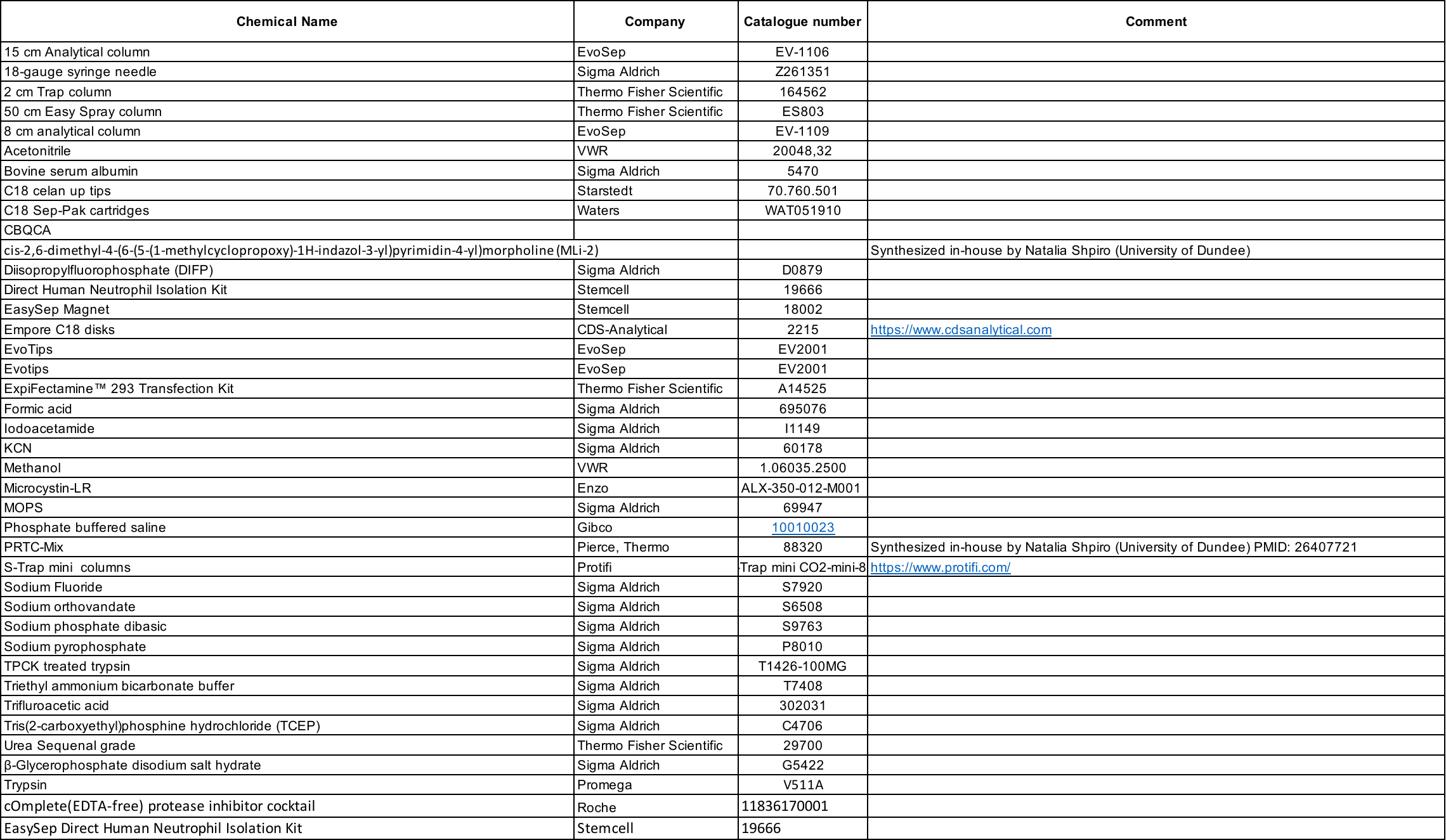

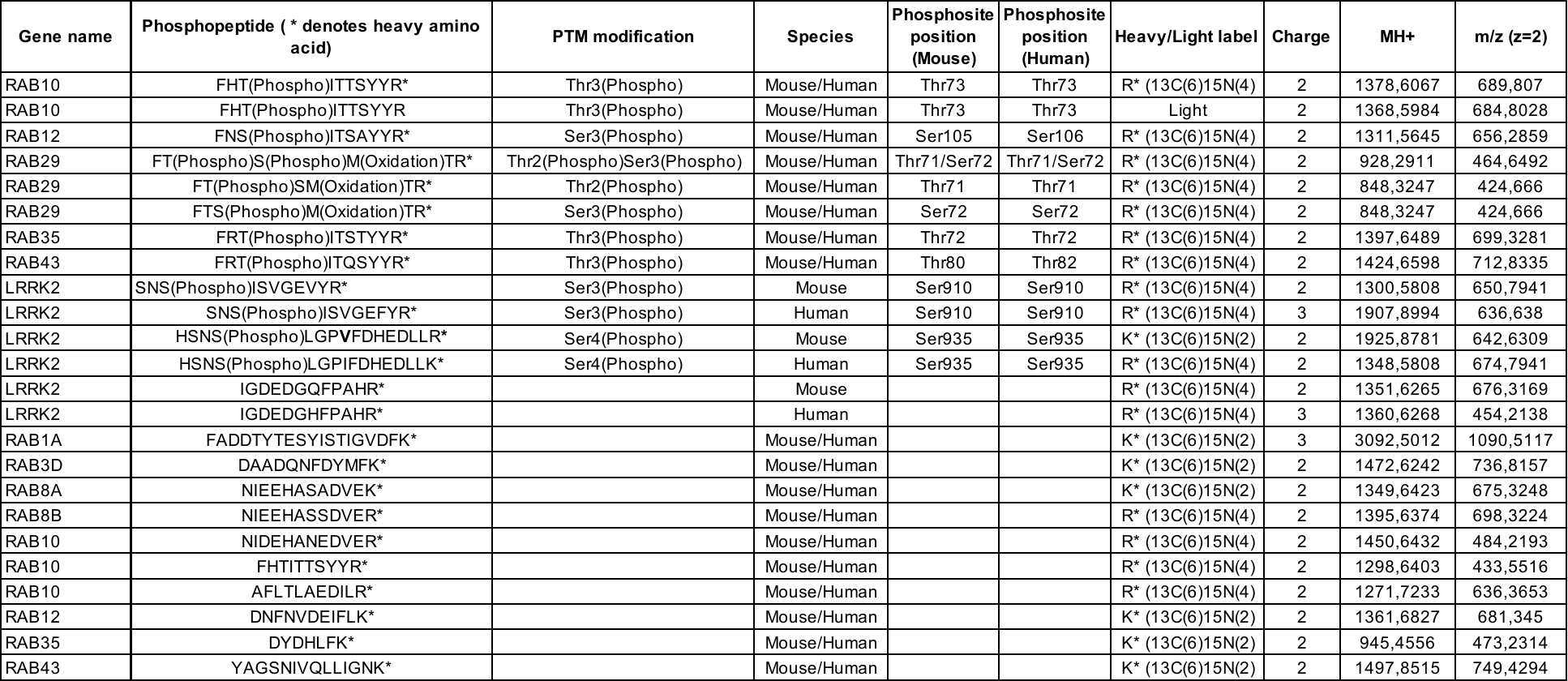

### Supplementary table S2

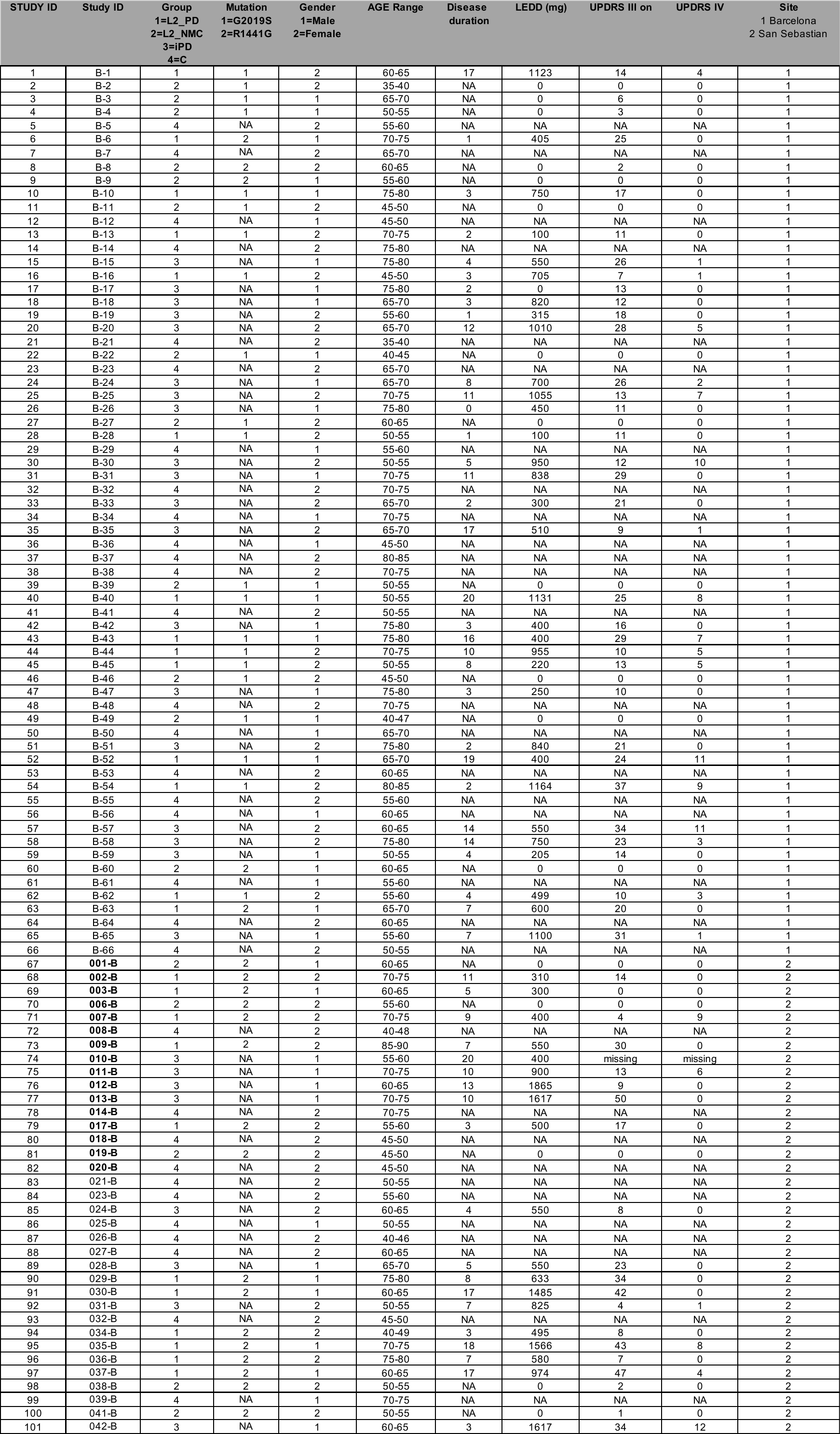

### Supplementary table S3

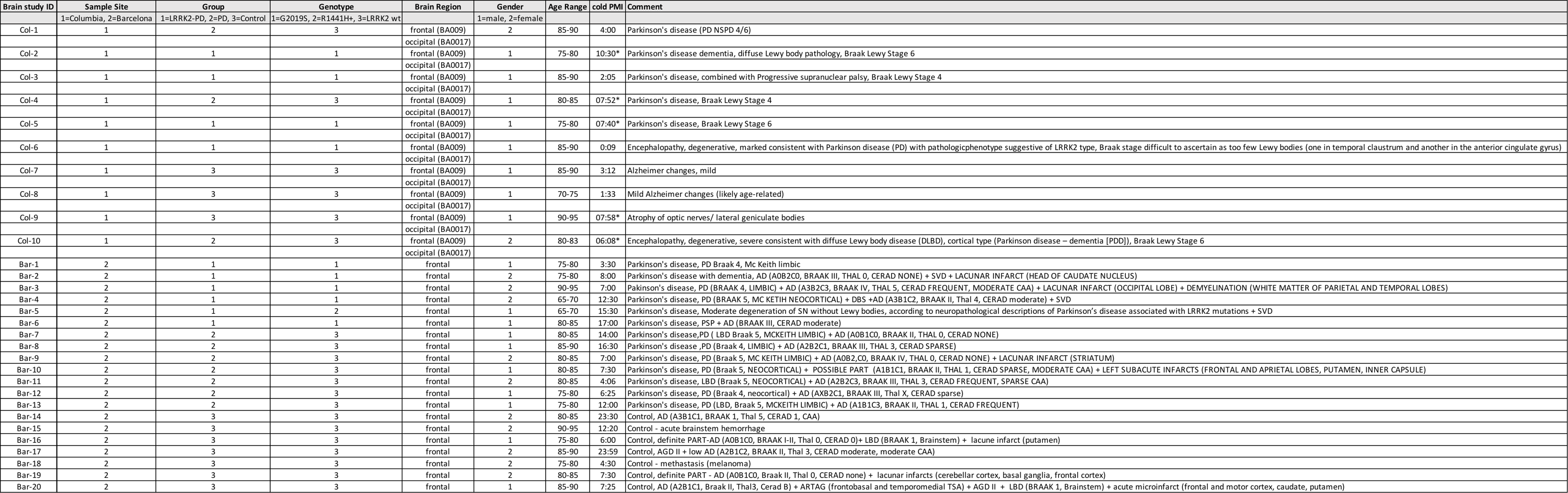
